# Distances from ligands as main predictive features for pathogenicity and functional effect of variants in NMDA receptors

**DOI:** 10.1101/2024.05.06.24306939

**Authors:** Ludovica Montanucci, Tobias Brünger, Nisha Bhattarai, Christian M Boßelmann, Sukhan Kim, James P. Allen, Jing Zhang, Chiara Klöckner, Piero Fariselli, Patrick May, Johannes R. Lemke, Scott J. Myers, Hongjie Yuan, Stephen F. Traynelis, Dennis Lal

## Abstract

Genetic variants in genes *GRIN1*, *GRIN2A*, *GRIN2B*, and *GRIN2D*, which encode subunits of the N-methyl-D-aspartate receptor (NMDAR), have been associated with severe and heterogeneous neurologic diseases. Missense variants in these genes can result in gain or loss of the NMDAR function, requiring opposite therapeutic treatments. Computational methods that predict pathogenicity and molecular functional effects are therefore crucial for accurate diagnosis and therapeutic applications. We assembled missense variants: 201 from patients, 631 from general population, and 159 characterized by electrophysiological readouts showing whether they can enhance or reduce the receptor function. This includes new functional data from 47 variants reported here, for the first time. We found that pathogenic/benign variants and variants that increase/decrease the channel function were distributed unevenly on the protein structure, with spatial proximity to ligands bound to the agonist and antagonist binding sites being key predictive features. Leveraging distances from ligands, we developed two independent machine learning-based predictors for NMDAR missense variants: a pathogenicity predictor which outperforms currently available predictors (AUC=0.945, MCC=0.726), and the first binary predictor of molecular function (increase or decrease) (AUC=0.809, MCC=0.523). Using these, we reclassified variants of uncertain significance in the ClinVar database and refined a previous genome-informed epidemiological model to estimate the birth incidence of molecular mechanism-defined GRIN disorders. Our findings demonstrate that distance from ligands is an important feature in NMDARs that can enhance variant pathogenicity prediction and enable functional prediction. Further studies with larger numbers of phenotypically and functionally characterized variants will enhance the potential clinical utility of this method.

## Introduction

Pathogenic variants in the GRIN family of genes encoding the N-methyl-D-aspartate receptor (NMDAR) subunits have been found in patients with various neuropsychiatric disorders, including autism spectrum disorders, epilepsy, intellectual disability, attention-deficit/hyperactivity disorder, and schizophrenia ^1–8^. NMDARs are tetrameric ligand-gated ion channels permeable to Na^+^, K^+^, and Ca^2+^, composed of two glycine-binding GluN1 subunits and two glutamate-binding GluN2 subunits, which can be a combination of any two of GluN2A, GluN2B, GluN2C, or GluN2D^9,10^. GluN1 subunits are encoded by the gene *GRIN1* and GluN2 subunits are encoded by the genes *GRIN2A, GRIN2B, GRIN2C*, and *GRIN2D*. Among the pathogenic variants identified in the GRIN gene family, those in *GRIN2A* (46%) and *GRIN2B* (38%) account for the vast majority, followed by *GRIN1* variants (14%)^11^. Variants in these genes have been associated with a spectrum of neurodevelopmental disorders^12^. For example, *GRIN1* and *GRIN2B* patients can present with mild or severe intellectual disabilities^1,3,13^. While some *GRIN2A* patients have severe intellectual disabilities, roughly half have no intellectual impairment^8,14^. Most patients with variants in *GRIN2A* have seizures whereas the majority of patients with variants in *GRIN2B* do not have seizures^4^. In addition, low muscle tone is rare among *GRIN2A* patients while common among patients involving other GRIN genes^15,16^. All GRIN patients present with speech impairment, even those without intellectual disabilities ^17^. *GRIN2D* patients appear to have the most severe phenotype, although there is not yet enough data to understand the full clinical spectrum^15,16^. More recently, patients with protein-truncating variants in *GRIN2A* have been associated with schizophrenia^18^ and are susceptible to seizures and delayed maturation of parvalbumin interneurons, both of which resolve after adolescence^19^.

NMDARs serve many cellular functions, including both pre-synaptically to influence neurotransmitter release and long-term plasticity^20–22^ and post-synaptically to mediate the slow component of postsynaptic currents and synaptic plasticity^23,24^. NMDARs function as signal coincidence detectors, as their activation requires changes in membrane potential to relieve the pore blocking by Mg^2+^ ions as well as the synaptic release of glutamate ^10,25,26^. Thus, NMDARs are precisely regulated by the binding of glutamate and its co-agonist glycine, of extracellular Mg^2+^ that blocks the channel, and of other endogenous extracellular modulators such as zinc ions (Zn^2+^)^27^. Genetic variants in GRIN genes can cause a heterogeneous spectrum of alterations of the NMDAR function, which can be grouped into two main types: gain of the NMDAR function (or gain-of-function effect or GoF) and partial or complete loss of the NMDAR function (or loss-of-function effect or LoF) ^28^. Because of the many functions and modulators of NMDARs, a large number of NMDAR-based molecules have been developed as therapeutic options designed to mitigate dysfunction of the glutamatergic system ^29,30^.

Variant interpretation in the GRIN genes, both for pathogenicity and molecular functions, is still challenging. Currently, more than 65% of missense variants in GRIN genes are classified as variant of unknown significance according to the ClinVar database (accessed July 2022)^31^. To improve variant interpretation, several exome-wide bioinformatic approaches have been developed that can identify clusters of patient variants or population variant depletion across genomic, protein sequences or 3D structures^32–37^. These methods and targeted clinical-genetic studies for *GRIN2A* and *GRIN2B* showed enrichment of pathogenic over population variants in several UniProt defined domains^13,37,38^. Although these approaches can identify important regions, they typically don’t explore the underlying structure-to-function relationship. However, in the absence of functional characterization of every possible genetic variant in NMDAR encoding genes, prediction models are needed to enable precision care since different disease mechanisms have contraindicated treatment needs^39–42^. For example, among patients with developmental and epileptic encephalopathy, those with a variants that cause gain of NMDAR function represent candidates for potential treatment with NMDAR blockers, such as memantine^43–45^ or GluN2B-selective inhibitors^42,46^, while those with complete or partial loss of the NMDAR function may potentially respond to positive allosteric modulators of the NMDAR^47–49^.

However, the functional consequences are not known for most of the variants. Electrophysiological studies that experimentally determine the molecular functional consequences introduced by missense variants are expensive and time-consuming, and it is difficult to envision how all the possible GRIN missense variants can be functionally assessed. Machine learning (ML)-based methods may be able to take advantage of the limited available experimental data to predict the molecular functional consequences of the NMDAR variants that have not been experimentally tested, as has been proven successful for example in voltage-gated potassium channels^50^ and sodium channels^51^. These methods are based on sequence and structural protein features, as discriminative gene-level and protein-level features have been found to be associated with the GoF/LoF effects of variants ^52–54^. To date, few predictors are available for variant functional effects (VPatho^55^, LoGoFunc https://www.biorxiv.org/content/10.1101/2022.06.08.495288v1.full.pdf) and none are specifically designed for GRIN variants. Here we seek to identify structural features of missense variants that are predictive of variant pathogenicity and of increased or decreased functional effects that are specific for the GRIN genes to develop a ML-based method to predict pathogenicity and Increased/Decreased consequences in GRIN genes. Therefore, we aggregated a unique set of 201 expert-curated patient variants and 631 population variants from the gnomAD database together with 159 functionally characterized missense variants from electrophysiological readouts across *GRIN1*, *GRIN2A* and *GRIN2B*. Some of these variants were functionally characterized in this study for the first time. Since previous work^12,56–60^ described how individual missense variants in NMDARs alter their ligand-induced regulation, we first sought whether spatial distance from agonists that are bound to their binding sites in the NMDAR protein structure correlates with pathogenicity and increased/decreased functional effects. With the identified distances from ligands and additional biophysical and evolutionary scores, we have built a ML-based predictor specifically trained to classify variants in GRIN genes as pathogenic or benign, and a ML-based predictor specifically trained to classify the molecular consequences of missense variants into increased or decreased effect, thus providing a valuable resource for clinical genetics.

## Material and Methods

### Clinical data set of GRIN missense variants

#### Clinical dataset

The variant data set comprises manually curated patient missense variants collected through patient registries. Clinical cases were collected using a REDCap survey with 391 fields on genetic and clinical data (REDCap version 10.9.3 https://www.project-redcap.org/) and offer the integration of retrospective and longitudinal data. All clinical cases were manually reviewed and classified in accordance with American College of Medical Genetics (ACMG) guidelines. These variants were found in the genes *GRIN1, GRIN2A*, and *GRIN2B*. Missense variants from the general population in the same genes were derived from the gnomAD database^61^. In order to map the variants on the NMDAR structure, we restricted our analysis to missense variants that were localized in the domains of the NMDAR structure that are atomically resolved in the PDB: the amino-terminal (ATD), agonist-binding (ABD), and transmembrane (TMD) domains. Missense variants that are localized in C-terminal domain (CTD) could not be considered as the CTD is not present in any resolved NMDAR structure. This comprises a set of 832 missense variants, of which 201 from patients and 631 from controls. The full list of the considered variants along with their clinical annotation is available and interactively accessible at https://GRIN-portal.broadinstitute.org and and also reported in Supplemental Table S1.

### Functional data set of GRIN missense variants

We identified 127 missense variants in *GRIN1*, *GRIN2A* and *GRIN2B* for which functional testing through electrophysiological and biochemical assays were published and completed to an extent to allow GoF and LoF determination by the criteria of Myers et al. 2023^28^ (Supplemental Table S2). We also identified additional variants in the literature for which some functional data exists, but which lack completion of all assays needed to classify by criteria described in Myers et al. (2023; Supplemental Table S3). Among these two sets of variants, we present new data allowing completion of functional and biochemical assessment of 34 known missense variants according to the criteria of Myers et al. (2023; Supplemental Table S4, S5). From these determinations, we categorized a total of 159 variants as either having an increased or decreased ion channel function of the response for our implementation here by evaluating the criteria described in Myers et al. (2023)^28^, omitting consideration of surface expression, since our algorithms explore receptor function as it relates to protein structure, not factors that influence trafficking. Thus, functional data from 159 variants is the starting point for our functional analyses.

#### Functional analysis of missense variants

cDNAs encoding human NMDAR subunits GluN1-1a (hereafter GluN1; GenBank accession codes: NP_015566), GluN2A (GenBank accession codes: NP_000824), and GluN2B (GenBank accession codes: NM_000825) were used and site-directed mutagenesis used to introduce human variants (QuikChange; Stratagene, La Jolla, CA, USA); all mutant cDNAs were verified by dideoxy DNA sequencing (Eurofins MWG Operon, Huntsville, AL, USA). The cDNA for WT and mutant NMDAR subunits was linearized using the appropriate restriction enzyme and cRNA was synthesized *in vitro* using the mMessage mMachine T7 kit (Ambion, Austin, TX, USA).

*Xenopus laevis* ovaries with unfertilized oocytes (Stage V-VI) were obtained from Xenopus One Inc (Dexter, MI, USA) and digested with Collagenase Type 4 (Worthington-Biochem, Lakewood, NJ, USA; 850 μg/ml, 15 ml for a half ovary) in Ca^2+^-free Barth’s solution that contained (in mM) 88 NaCl, 2.4 NaHCO_3_, 1 KCl, 0.82. maintained at 16°C and injected with cRNA encoding either WT or variant NMDAR subunits (GluN1:GluN2A or GluN2B ratio 1:2, 5-10 ng total in 50 nl of RNAase-free water per oocyte). Injected oocytes were maintained in normal Barth’s solution at 16-19°C.

Two-electrode voltage clamp (TEVC) current recordings from Xenopus oocytes expressing NMDARs were performed as previously described ^62,63^. Oocytes were transferred to a recording chamber and were perfused with extracellular recording solution composed of (in mM) 90 NaCl, 1 KCl, 0.5 BaCl_2_, 10 HEPES, and 0.01 EDTA (23°C, pH 7.4 with NaOH, EDTA omitted in experiment measuring Mg^2+^IC_50_). Current responses to glutamate and glycine were recorded under voltage clamp at a holding potential of −40 mV; current and voltage electrodes were filled with 3 and 0.3 M KCl, respectively. Maximally effective concentrations of agonists (100 μM glutamate and 100 μM glycine) were used unless stated otherwise. The reagent 2-aminoethyl methanethiol sulfonate hydrobromide (MTSEA; Toronto Research Chemicals, Ontario, Canada) was made fresh and used within 30 min.

HEK293 cells (ATCC CRL-1573) were plated on glass coverslips coated with 0.1 mg/ml poly-D-lysine and maintained in Dulbecco’s modified Eagle medium (DMEM) with 10% fetal bovine serum and 10 U/ml streptomycin at 37°C (5% CO_2_). The cells were transfected with cDNA encoding GluN1, GluN2A, and eGFP at a ratio of 1:1:5, or GluN1, GluN2B, and eGFP at a ratio of 1:1:3 using the calcium phosphate method^62^. 12-24 hrs post transfection the cells were transferred to the recording chamber and perfused with recording solution composed of (in mM) 150 NaCl, 3 KCl, 1.0 CaCl_2_, 10 HEPES, 0.01 EDTA, and 2.10 D-mannitol (the pH was adjusted to 7.4 with NaOH). 3-5 MΩ fire-polished patch electrodes were made from thin-walled micropipettes (TW150F-4, World Precision Instruments, Sarasota, FL, USA) and filled with internal solution composed of (in mM) 110 D-gluconate, 110 CsOH, 30 CsCl, 5 HEPES, 4 NaCl, 0.5 CaCl_2_, 2 MgCl_2_, 5 BAPTA, 2 NaATP and 0.3 NaGTP (the pH was adjusted to 7.4 with CsOH; osmolality was 300-305 mOsmol/kg). Whole cell currents in response to application of 1.0 mM glutamate and 100 μM glycine (V_HOLD_ −60 mV, 23°C) were recorded by an Axopatch 200B patch-clamp amplifier (Molecular Devices, Union City, CA, USA). The current responses were filtered at 8 kHz (−3 dB) with an 8-pole Bessel filter (Frequency Devices) and digitized at 20 kHz on a Digidata 1440A system controlled by Clampe× 10.3 (Molecular Devices). The position of double-barreled theta-glass tubing was controlled by a piezoelectric translator to obtain rapid solution exchange (Burleigh Instruments, Newton, NJ, USA). Large current responses were corrected off-line for series resistance errors^64^.

To measure receptor surface expression, HEK 293 cells grown in 96-well plates (50,000 cells/well) were transfected with cDNA encoding beta-lactamase (β-lac) fused in frame to the N-terminus of GluN1 with WT or mutant GluN2, or similarly constructed β-lac-GluN2 with WT or mutant GluN1 using Fugene6 (Promega, Madison, WI)^56^. Wells treated with Fugene6 alone without cDNA were used to determine background absorbance. NMDAR antagonists (200 μM DL-APV and 200 μM 7-CKA) were added to cultures when transfected. Six wells were transfected for each variant to determine surface and total protein levels (3 wells each). After 24 hours, cells were rinsed with Hank’s Balanced Salt Solution (HBSS) that was composed of (in mM) 140 NaCl, 5 KCl, 0.3 Na_2_HPO_4_, 0.4 KH_2_PO_4_, 6 glucose, 4 NaHCO_3_ with 10 mM HEPES added. Subsequently, 100 μl of a 100 μM nitrocefin (Millipore, Burlington, MA, USA) solution in HBSS plus HEPES was added to each of the wells and extracellular enzymatic activity was determined. The cells in the three wells were lysed by 50 μl H_2_O (30 min) prior to the addition of 50 μl of 200 μM nitrocefin to determine total enzymatic activity. The absorbance at 468 nm was read every min for 30 min at 30°C, and the rate of increase in absorbance was determined from the slope of a linear fit to the data.

### Structural localization of variants

In order to investigate correlations between functional effects (pathogenicity and molecular function) and structural features of NMDAR missense variants, we localized each variant onto the 3-dimensional (3D) protein structure using SIFTS tools^65^ to cross reference amino acid positions between protein sequences and protein structures. We used two crystal structures of the human NMDAR available in the PDB: 7EU7^66^ (3.50 Å of resolution) which comprises two *GRIN1*-encoded and two *GRIN2A*-encoded chains, and 7EU8^66^ (4.07 Å of resolution) which comprises two *GRIN1*-encoded chains and two *GRIN2B*-encoded chains. We used chain A and B of 7EU7^66^ to localize *GRIN1* and *GRIN2A* variants respectively, and chain B of 7EU8 to localize *GRIN2B* variants. Of the variants in our collected clinical dataset, 832 could be mapped onto the heterotetrametric protein structure. Of these, 201 are from patients and 631 from population.

### Computation of distance features for missense variants

For each variant that could be mapped onto the 3D structure, we computed features that synthesize information about their position in the 3D structure with respect to the functionally important regions of the protein complex. In particular, we computed their distances from: the membrane center, the pore axis, and the four ligands known to primarily regulate NMDAR function (glutamate, glycine, magnesium ion Mg^2+^, and zinc ion Zn^2+^). We used the protein complex 7EU7^66^ to compute distances for *GRIN1* and *GRIN2* variants and 7EU8^66^ for *GRIN2B* variants. We annotated the membrane through the PPM server (https://opm.phar.umich.edu/ppm_server, version PPM 2.0)^67^ and calculated the minimum distance of the wild-type residue of each variant from the membrane center as defined by the PPM server. We annotated the pore using the Mole2.5 webserver^68^ (https://mole.upol.cz/, access 24.03.2022) and calculated the minimum distance of the wildtype residue of each variant from the pore axis using R-script and the “bio3D” package^69^. To calculate the minimum distance of the wildtype residue of each variant from the ligands (glutamate, glycine, Mg^2+^, Zn^2+^) we first mapped these ligands to the corresponding reference protein structures 7EU7 and 7EU8. Two ligands, glutamate and glycine, were already crystallized in the protein complex 7EU7. To map these two ligands to the 7EU8 protein complex, we performed a structural alignment of the two protein complexes (7EU7 and 7EU8) using mTM-align^70^ (root mean square error (RMSD): 3.22Å). To place Zn^2+^, we carried out two structural alignments of the amino terminal domain of the NMDAR crystallized with two Zn^2+^ ions bound to GluN2A (PDB-ID: 5TQ2, resolution of 3.29 Å) with 7EU7^66^ and 7EU8^66^, respectively. The structural alignments were performed with the mTM-align^70^ program (RMSD_7EU7_ = 3.01Å, RMSD_7EU8_ = 2.22Å). Since no NMDAR structure has been crystallized with Mg^2+^ inside yet, we calculated the estimated the location of the Mg^2+^ ions by calculating the center of the described magnesium binding residues (N615 *GRIN2A*, N616 *GRIN2B* and N616 *GRIN1)*^2,71^. For each missense variant, we then calculated the minimum distance of its wild-type residue from each ligand. The distance calculation was restricted to residues located in the same domain. Domain annotations are detailed in Supplementary Table S1 and are: amino terminal domain (ATD), agonist binding domain (ABD), ABD-TMB linkers (S1 and S2), and transmembrane domain (TMD) comprising M1, M2, M3, and M4 helices. All distances were computed considering all atoms. The considered distance features are tabulated in Table 1.

**Table 1.**
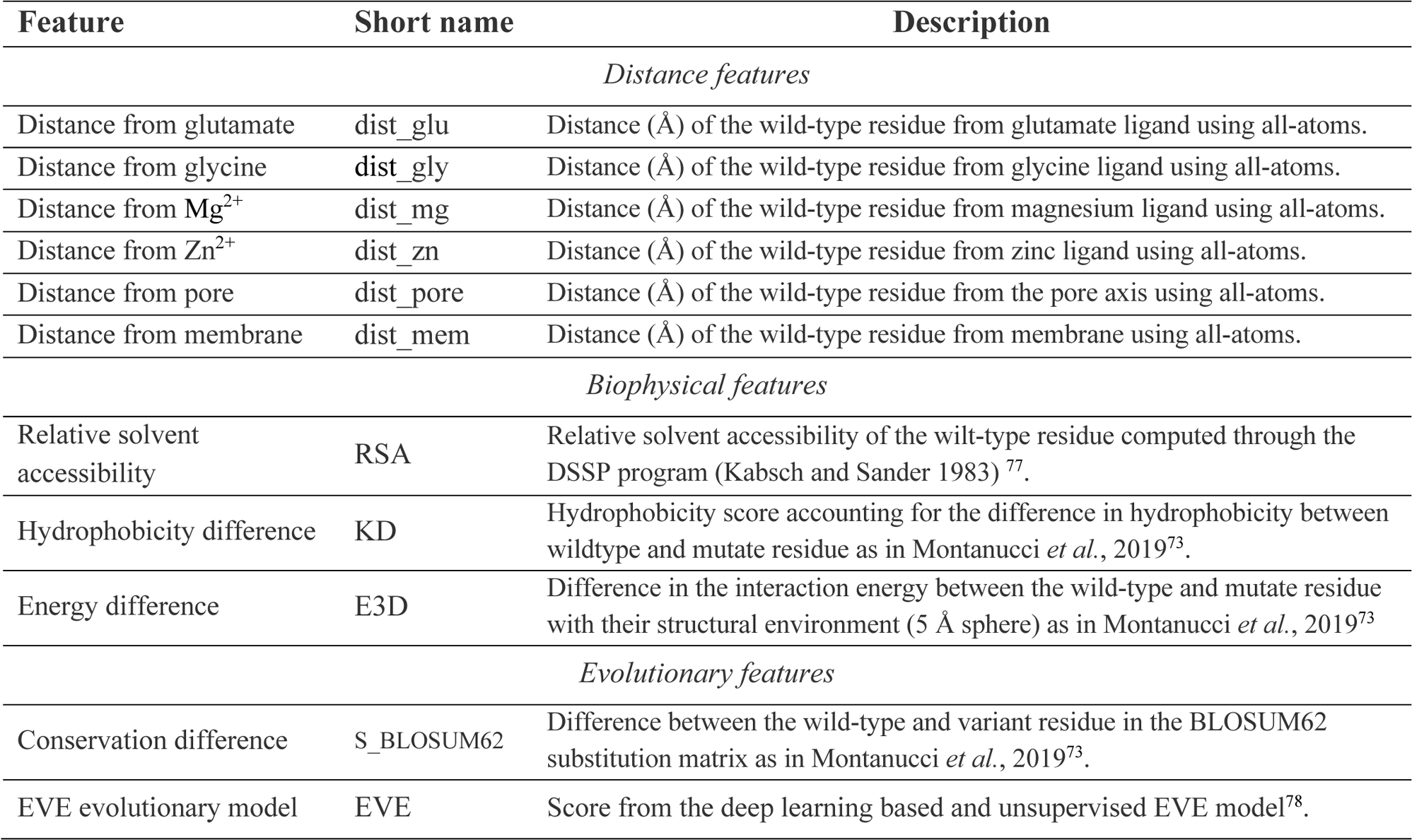
List of the considered distance, biophysical, and evolutionary features for missense variants.

### Computation of biophysical and evolutionary features for missense variants

In addition to distance features, we also computed biophysical and evolutionary scores. For each variant, we computed three biophysical features: the relative solvent accessibility of the wild-type residue computed through the DSSP program, the difference in hydrophobicity between the wild-type and the substituted residues according to the Kyte-Dolittle scale and the difference in the interaction energy (computed as the Bastolla-Vendruscolo statistical potential^72^) between the wild-type and mutate residue with their structural environment (5 Å sphere centered in the mutated residue). These two latter scores have been weighted through the sequence profile as in Montanucci *et al.*, 2022^69^. The biophysical features are listed in Table 1. In addition to biophysical scores, we computed, for each variant in our clinical and functional dataset the BLOSUM62^75^ and EVE^76^ evolutionary conservation scores. These scores for each variant are tabulated in Supplementary Table 1.

### Enrichment analysis

We performed an enrichment analysis to reveal the distribution of pathogenic and population variants and of variants with increased and decreased functional consequence on the NMDAR structure. To perform the enrichment analysis, we applied the Wilcoxon rank sum test to investigate differences in patient and population variants and their 3D distances from functionally relevant protein region sites (pore axis and membrane) and the four selected ligands. Similarly, we tested differences for variants associated with an increased or decreased functional effect. We applied Bonferroni correction to account for multiple testing.

### Developing of ML-based binary classifiers for pathogenicity and functional prediction

#### Pathogenicity predictor

In order to develop a binary ML-based predictor which classifies missense variants in the GRIN genes as benign or pathogenic, we trained each of four binary predictors, each one on a different set of features, listed in Table 2. PP-dist, PP-evo, PP-biophys trained on only distances from ligands and pore axis, evolutionary scores, biophysical features, respectively. Finally, PP-dist&evo is trained on the combination of features of the two best performing predictors, which are distances and evolutionary features. All the predictors have been trained on the subset of the clinical dataset comprising variants that could be mapped on the 3D structure. A 5-fold cross-validation procedure was applied.

**Table 2.** Description of the input features for each pathogenicity and functional effect predictor.

| Predictor | Input Features* |
| --- | --- |
| *Pathogenicity Predictor (PP)* |  |
| PP-dist | dist_glu, dist_gly, dist_mg, dist_zn, dist_pore |
| PP-evo | S_BLOSUM62, EVE |
| PP-biophys | RSA, KD, E3D |
| PP-dist&evo | dist_glu, dist_gly, dist_mg, dist_zn, dist_pore, S_BLOSUM62 |
| *Functional (Increase/Decrease) Predictor (FP)* |  |
| FP-dist | dist_glu, dist_gly, dist_mg, dist_zn |
| FP-evo | S_BLOSUM62, EVE |
| FP-biophys | RSA, KD, E3D |
| FP-dist&evo | dist_glu, dist_gly, dist_mg, dist_zn, S_BLOSUM62, EVE |

\*Short names of the biophysical features are derived from Table 1.

#### Functional predictor

In order to develop a binary ML-based predictor which classifies missense variants in the GRIN genes as increasing or decreasing functional effect, we trained four binary predictors, each one trained on a different feature set, listed in Table 2. FP-dist is trained on distances from the four ligands: glutamate, glycine, Mg^2+^, and Zn^2+^; FP-evo is trained on evolutionary scores; FP-biophys is trained on the three biophysical features of the variant. Finally, FP-dist&evo is trained on distance and evolutionary features. The predictors have been trained on the functional dataset comprised of 159 variants, of which 89 with increased and 71 with decreased functional effect (see Supplementary Table 2). Details about machine learning implementation, cross-validation and indexes to evaluate performances are described in the Supplementary Notes. In order to compare the performance with existing variant functional effects predictors, we retrieved the prediction of our 159 functionally characterized variants of LoGoFunc. At the moment of submission VPatho was not available.

## Results

### Spatial proximity to ligands and pore is different in pathogenic versus benign variants

Here we asked whether the spatial distance from important ligands, such as glutamate, glycine, magnesium and Zn^2+^ and functional important sites such as the pore-axis correlate with variant pathogenicity in the *GRIN1*, *GRIN2A* and *GRIN2B* determined using ACMG criteria. We calculated the 3D distances of the 832 variants that could be mapped on the deposited NMDAR structures from the pore, the membrane and the four ligands that regulate the NMDAR activity and we compared the residue distances from these sites between the two groups of variants, an expert-curated set of patient variants (see Supplemental Tables S2-6).

Compared to the spatial distribution of population variants, patient variants where closer to the pore (Population variants *_Median distance_* = 35 Å, Patient variants *_Median distance_* = 18 Å, *P* =6.8e-45, Wilcoxon rank sum test, Figure 1B) and to the closest ligand (Population variants *_Median distance_* = 23 Å, Patient variants *_Median distance_* = 17 Å, *P* = 3.7e-15, Wilcoxon rank sum test, Figure 1C). When performing the same enrichment analysis for each domain separately, we found that in the agonist binding domain, *GRIN2A* and *GRIN2B* patient variants in the agonist binding domain are located closer to glutamate, which is bound in the cleft of the bilobed agonist binding domain, compared to population variants (*P_GRIN2A_*= 4.3e-14, and *P_GRIN2B_*= 4.1e-03, Figure 2A), while no significant difference in proximity to glycine was found for variants in *GRIN1*. In the amino terminal domain, no significant difference is found in proximity to ligands between patient variants. However, this result could be due to the small number of population variants in this domain. In the transmembrane binding site, we observed that patient variants are closer to the Mg^2+^ binding site compared to population variants in *GRIN2A* (P = 2.6e-02, Figure2D).

**Figure 1.**
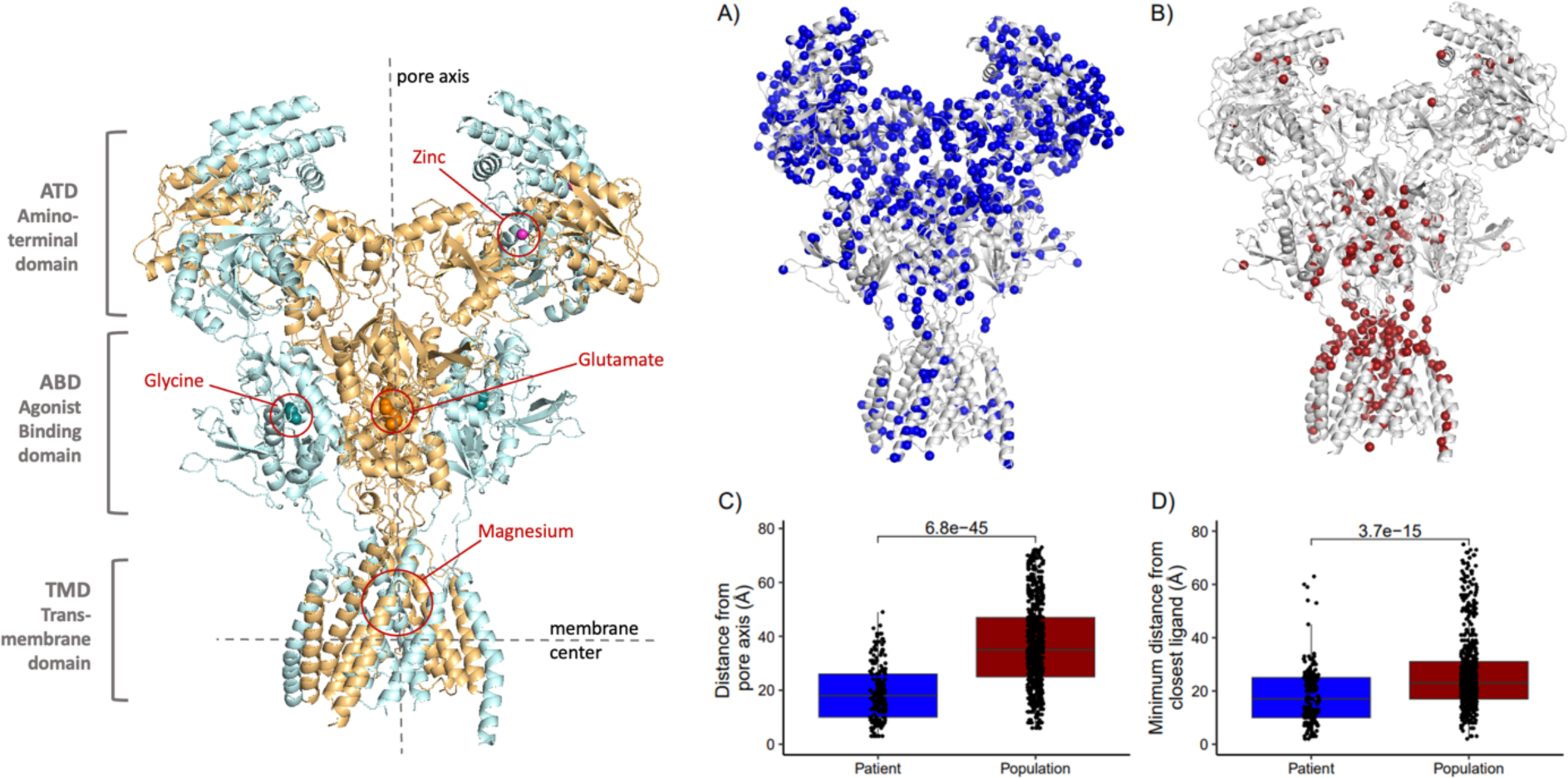
Variant distance from the pore axis and from ligands significantly differs between patient and population variants in the GRIN-genes. Left image: GluN1-GluN2A heterotetramer protein complex (PDB-ID: 7eu7). GluN1 subunits are colored in cyan, GluN2A subunits are colored in orange. The four ligand binding sites are highlighted. **A)** NMDAR structure with the 631 population variants of this study highlighted in blue. **B)** NMDAR structure with the 201 patient variants of this study highlighted in red. **C)** Boxplot of the distance from the pore axis for 201 patients and 631 population missense variants in *GRIN1*, *GRIN2A*, and *GRIN2B*. **D)** Boxplot of the minimum distance from the closest ligand (glutamate, glycine, Zn^2+^and Mg^2+^) for 203 patients and 631 population missense variants in *GRIN1*, *GRIN2A*, and *GRIN2B*. To quantify the differences in the distances to the ligands and the pore axis we performed the Wilcoxon-rank sum test and corrected for eight tests using Bonferroni correction.

**Figure 2.**
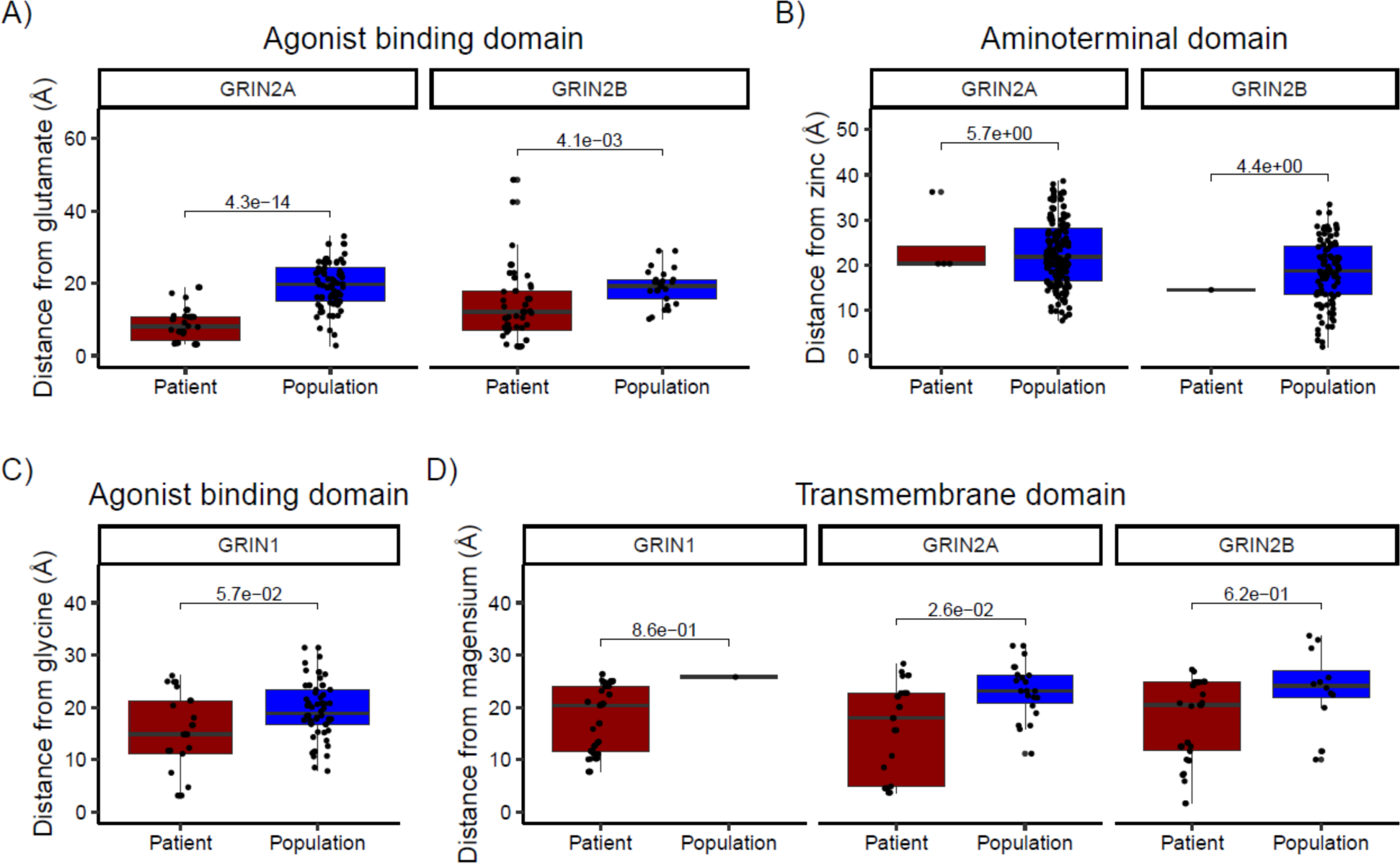
Variant distance from NMDAR ligands significantly differs between patient and population variants in the GRIN-genes. Boxplot of the distances of patient and population variants from four different NMDAR ligands: **A)** glutamate, **B)** Zn^2+^, **C)** glycine, and **D)** Mg^2+^. Only variants that are located in the same domain where the ligand is bound are shown and the distance is only computed for the protein subunits that are involved into the ligand binding. **A)** Distances from glutamate of patient and population variants that are located in the agonist binding domain (ABD). **B)** Distances from the Zn^2+^ of patient and population variants that are located in the amino terminal domain. **C)** Distances from glycine of patient and population variants that are located in the agonist binding domain (ABD). **D)** Distances from Mg^2+^ of patient and population variants that are located in the transmembrane domain (TMD). To quantify the differences in the distances to the ligands, we performed the Wilcoxon-rank sum test and corrected for eight test using Bonferroni correction.

### Machine learning method based on ligand and pore proximity to predict variant pathogenicity

Given the strong variant position to pathogenicity associations that we observed, we explored whether our generated distance features could be used to develop a method for the prediction of variant pathogenicity in the GRIN genes using our collected clinical dataset of 201 patient and 631 population variants. We developed a ML-based method, PP-dist, based on only distance features (distance from pore axis, glutamate, glycine, Mg^2+^ion and Zn^2+^) to classify a GRIN variant as pathogenic or benign. As a comparison, we trained two additional pathogenicity predictors on the same clinical dataset, PP-evo trained with only evolutionary features, and PP-biophys trained on biophysical properties of the amino-acid substitution (see Table 2 for input features of each predictor). The binary classifier based on only distances from ligands, PP-dist, reaches high prediction performances, with an overall accuracy of 0.892, an area under the ROC curve (AUC) of 0.9237 and a Matthews correlation coefficient (MCC) of 0.698 (see Table 3 and Figure 3). The predictive power of these distances is therefore very high, considering that these input features are based on only the position of the substituted residue and do not contain any information on the properties of the alternative residue. This indicates that the distance from pore and ligands, are major features in determining pathogenicity of variants in the GRIN genes. While PP-biophys shows a low MCC of 0.156 indicative of a poor predictor, PP-evo shows a high MCC (0.534) and an overall accuracy of 0.832 (Table 3 and Figure 3). This suggests that biophysical features provide much less information about variant pathogenicity than evolutionary scores. When a last predictor PP-dist&evo was trained with features from the two classes of distances from pore and ligands and evolutionary, the prediction performances improved by 1% in respect to the individual predictors PP-dist and PP-evo, reaching an overall accuracy of 0.903, a MCC of 0.726 and an AUC of 0.945.

**Table 3.**
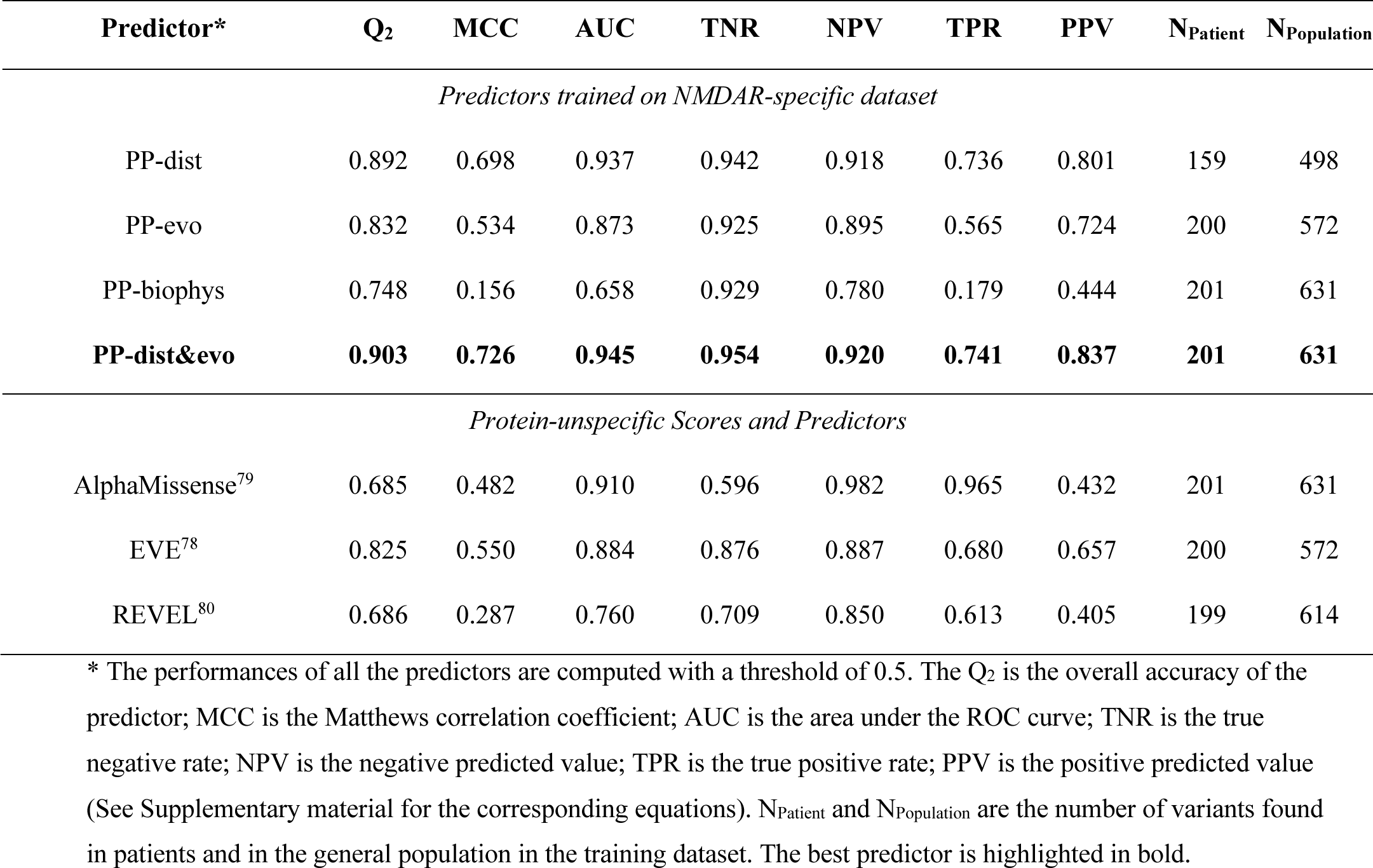
Performances of pathogenicity predictors on 838 variants in the NMDAR.

**Figure 3.**
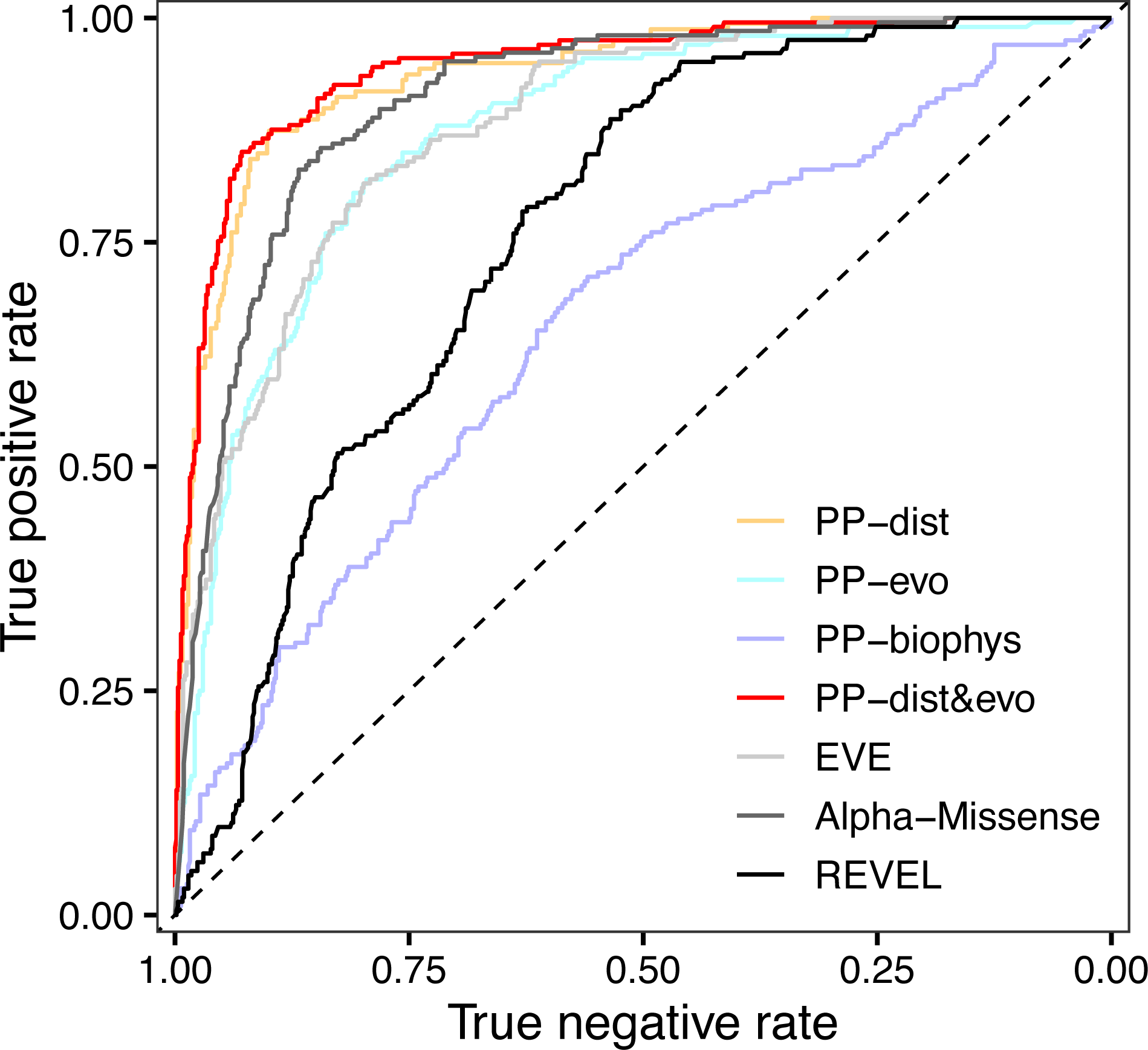
ROC curve for the ML-based pathogenicity predictors (PP-dist, PP-evo, PP-biophys and PP-dist&evo) and the three additional pathogenicity predictors and scores (AlphaMissense, EVE, and REVEL) on the dataset composed of the 832 GRIN variants of the clinical dataset.

To compare our developed predictors with existing methods, we also report the performances of other three among the main pathogenicity scores and predictors on the clinical dataset of this study (see Table 3 and Figure 3). This comparison shows that the simple distance measurements from ligands and pore allow a prediction accuracy that is ∼10% higher than the best non-gen-specific pathogenicity predictor.

### Validation of the pathogenicity predictor and pathogenicity prediction for variants of uncertain significance

So far, we only used expert curated patient variants. As a further validation, we applied our best pathogenicity predictor, PP-dist&evo, to classify variants that were in ClinVar (July 2022) and, at the same time, were not part of our expert curated dataset (for prediction scores see Supplementary Table 1). The total number of ClinVar variants that were not in our clinical dataset and were classified as (likely-) benign and (likely-) pathogenic and located in our 75% most confident class assignments is 44. Of these variants, 39 were correctly classified by our method, reaching a prediction accuracy of 0.89 (Figure 4A). We then used our best model, PP-dist&evo, to classify 100 ClinVar variants that were of uncertain or conflicting significance (VUS). Predictions for VUS were distributed across the whole spectrum of the pathogenicity score (Figure 4B). We reclassified 95 VUS that were assigned with a prediction score within the 75% most confident class assignments. Out of these 95 VUS, we predicted 19% (n = 18 VUS) as pathogenic and 81% (n = 77 VUS) as benign.

**Figure 4.**
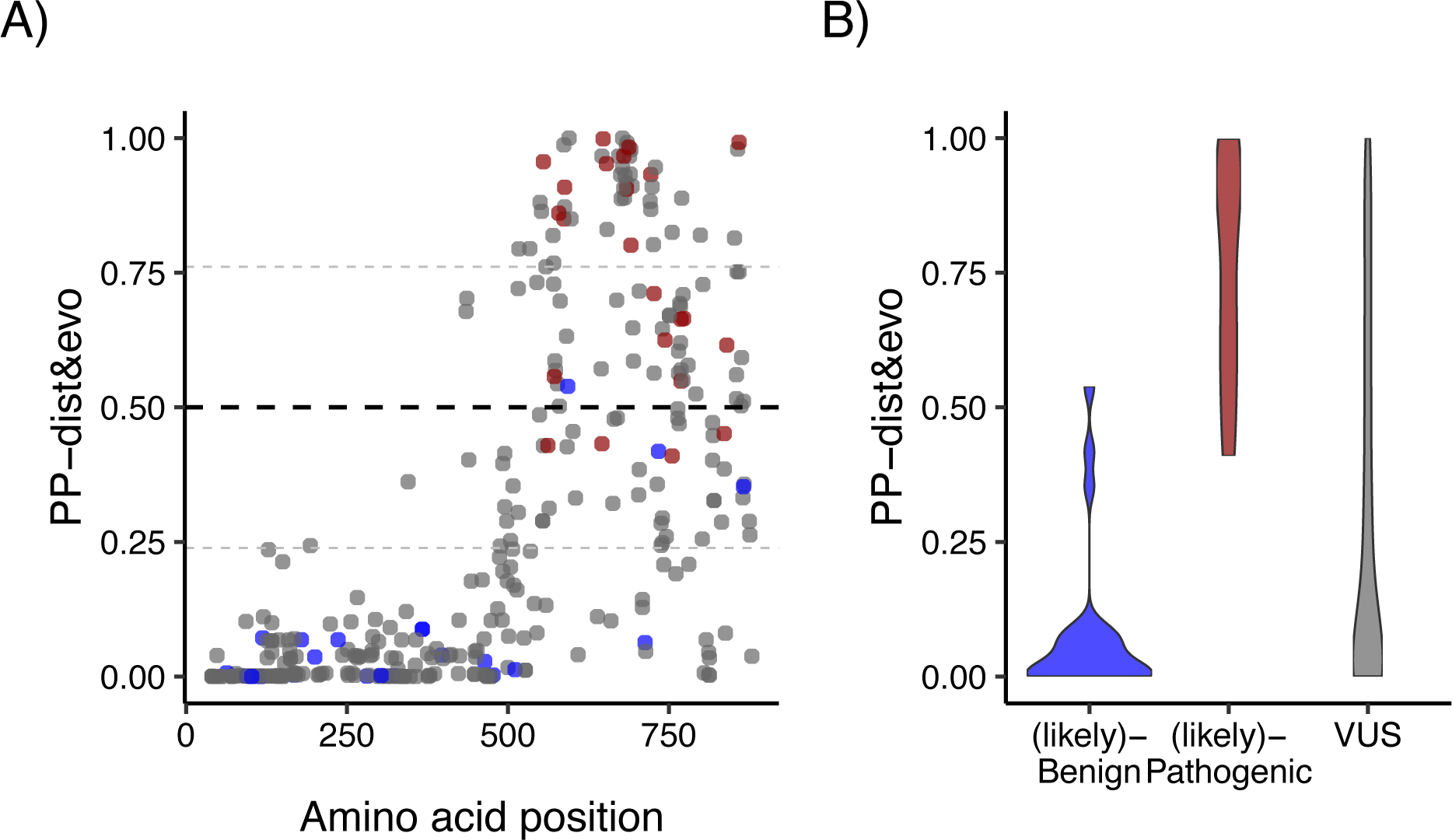
Pathogenicity prediction for ClinVar variants that were not part of our clinical dataset. **A)** Pathogenicity prediction for ClinVar variants that were not comprised in our clinical dataset. Prediction scores >0.5 indicate pathogenic variants, while predictor scores <0.5 indicate benign variants. The small grey line above and below 0.5 corresponds to the 75% most confident class assignments (over all possible amino acid variants). The prediction has been done with the best pathogenicity prediction method, PP-dist&evo. Prediction scores are displayed along the multiple sequence alignment of *GRIN1, GRIN2A & GRIN2B* (see alignment in the Supplementary Table 4). Blue, red, and grey dots correspond to ClinVar benign, pathogenic and VUS variants, respectively. **B)** Distribution of our prediction scores for ClinVar variants classified as benign, pathogenic and variants of unknown significance (VUS).

### Spatial proximity to ligands and pore is different in variants which cause an increased versus decreased functional consequence

After exploring the relationship between residue localization on the 3D-structure and pathogenicity of variants in the GRIN genes, we explored the relationship between the variant localization on the 3D-structure and the differential functional outcomes of the GRIN variants. We used data from 159 variants combining data from the peer reviewed literature and 47 variants generated in this study (Supplementary Table 2). In Figure 5 the boxplot of the distance of variants with different molecular effect is shown. Variants in *GRIN2A* and *GRIN2B* with decreasing effect are significantly closest (*P*=7.2e-3) to the glutamate in respect to variants with an increasing effect. Conversely, variants with increasing effect are significantly closest (*P*=4.3e-2) to Mg^2+^ in respect to variants with a decreasing effect. We evaluated function based on potency measurements for glutamate, glycine, Mg^2+^, and Zn^2+^. Because there is no reason a priori to assume that distance to the pore or agonist binding site will necessarily correlate with trafficking, which involves other parts of the receptor, we did not include an assessment of surface expression. This means that our categorization as Increasing and Decreasing function will not necessarily predict patient Gain-or Loss-of-Function as defined by Myers et al. (2023)^28^. Rather, this predictor will indicate whether a missense variant is likely to alter function of a receptor once it reaches the cell surface. In addition, for a subset of 69 variants, we did not have a measure of variant actions on deactivation time course, however this correlates with agonist EC_50_ and thus this effect is in part captured by our measure of potency in these variants (Xu et al., 2024) ^81,82^. We aggregated electrophysiological readouts from published and newly recorded data (Supplemental Table S1-3) in this study for 159 variants and classified all variants according to their molecular effect as described in the methods. We found the number of variants could be categorized as follows: N_Increase_ = 89, N_Decrease_ = 71. The results for this set of variants are summarized in Supplementary Tables S2-5. We observed that proximity to the pore, and in particular to the Mg^2+^ binding sites are associated with variants that “increased” function effects and depleted for variants that “decreased” function in all the GRIN genes (Figure 5D). In *GRIN2A* (P-value = 0.01), variants that “decreased” function are associated with close proximity to glutamate binding sites, compared to variants that “increased” function (Figure 5A).

**Figure 5:**
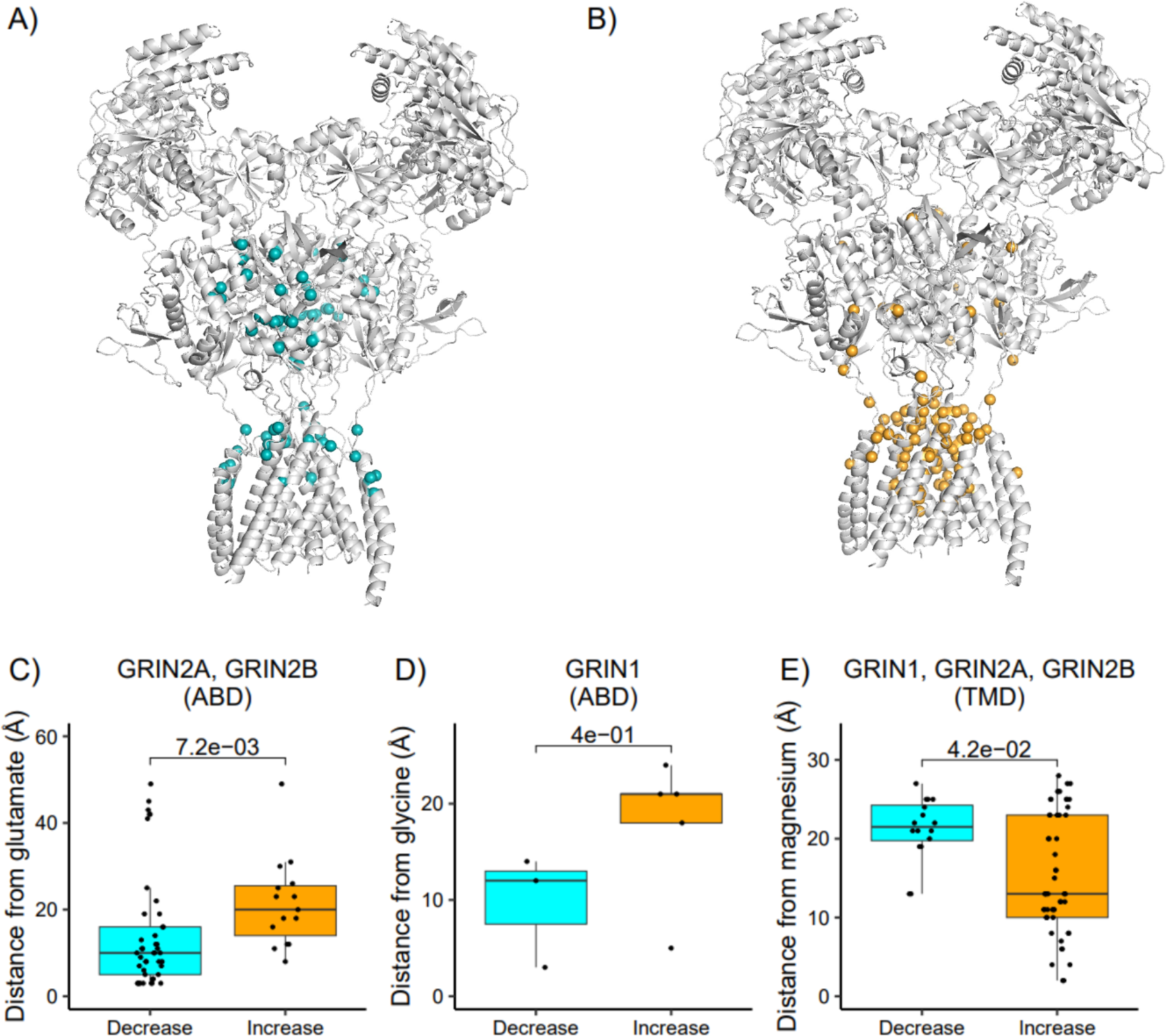
Variant distances from NMDAR ligands significantly differ between variants with different electrophysiological readouts in the GRIN-genes. In plot **A)** and **B)** variants whose molecular effect was classified into either decreasing **(A)** or increasing **(B)** effect are shown on the NMDA structure. In plots **C)**, **D)** and **E)** the boxplot of the distances between the variant and the ligands for the variants with different molecular effect are shown. Only variants that are located in the domain where the target ligand binds are considered. **C)** distance from glutamate, **D)** distance from glycine, and E) distance from Mg^2+^.

### ML-based method using ligand and pore proximity to predict functional effect of variants

With a similar procedure as that for pathogenicity predictor, we independently developed four different binary classifiers to predict the functional consequences of variants in *GRIN1*, *GRIN2A* and *GRIN2B* described in Table 2. All performances are shown in Table 4 and Figure 6. While the predictors based on only evolutionary or biophysical features (FP-evo and FP-biophys, respectively) are poor predictors (with MCC of 0.269 and 0.404, respectively), the FP-dist predictor, based only on the 3D distances of variants from each of the four considered ligands, reaches an overall accuracy of 0.740 and a MCC of 0.482. This shows here, for the first time, that distances of the variant from ligands contain information about the direction (“increasing” or “decreasing” activity) of the functional effect of the variant and that can be used for functional effect prediction.

**Table 4.** Performances of the binary predictors for the functional effect of variants (either increased or decreased function) on the data set of 159 experimentally characterized NMDAR variants.

| Predictor | Q<sub>2</sub> | MCC | AUC | TNR | NPV | TPR | PPV | N<sub>increase</sub> | N<sub>decrease</sub> |
| --- | --- | --- | --- | --- | --- | --- | --- | --- | --- |
| *Functional (Increased/Decreased function) predictors specifically trained on GRIN variants* |  |  |  |  |  |  |  |  |  |
| FP-dist | 0.740 | 0.482 | 0.800 | 0.644 | 0.776 | 0.828 | 0.716 | 64 | 59 |
| FP-evo | 0.6841 | 0.269 | 0.635 | 0.551 | 0.613 | 0.714 | 0.659 | 84 | 69 |
| FP-biophys | 0.703 | 0.404 | 0.740 | 0.500 | 0.761 | 0.871 | 0.679 | 85 | 70 |
| **FP-dist&evo** | **0.765** | **0.523** | **0.809** | **0.667** | **0.780** | **0.845** | **0.755** | **84** | **69** |
| *LoGoFunc predictor on the data set of 159 functionally tested GRIN variants* |  |  |  |  |  |  |  |  |  |
| LoGoFunc | 0.522 | 0.217 | 0.510 | 0.971 | 0.482 | 0.161 | 0.875 | 70 | 87 |
| *AlphaMissense evolutionary score on the data set of 159 functionally tested GRIN variants* |  |  |  |  |  |  |  |  |  |
| AlphaMissense | 0.491 | 0.193 | 0.651 | 1.000 | 0.467 | 0.080 | 1.000 | 71 | 88 |

The Q<sub>2</sub> is the overall accuracy of the predictor; MCC is the Matthews correlation coefficient; AUC is the area under the ROC curve; TNR is the true negative rate; NPV is the negative predicted value; TPR is the true positive rate; PPV is the positive predicted value (see Supplementary Material for details). N<sub>increase</sub> and N<sub>decrease</sub> are the number of variants experimentally determined as increased/decreased function in the training dataset for which all the features for each predictor were available. The best predictor is highlighted in bold.

**Figure 6.**
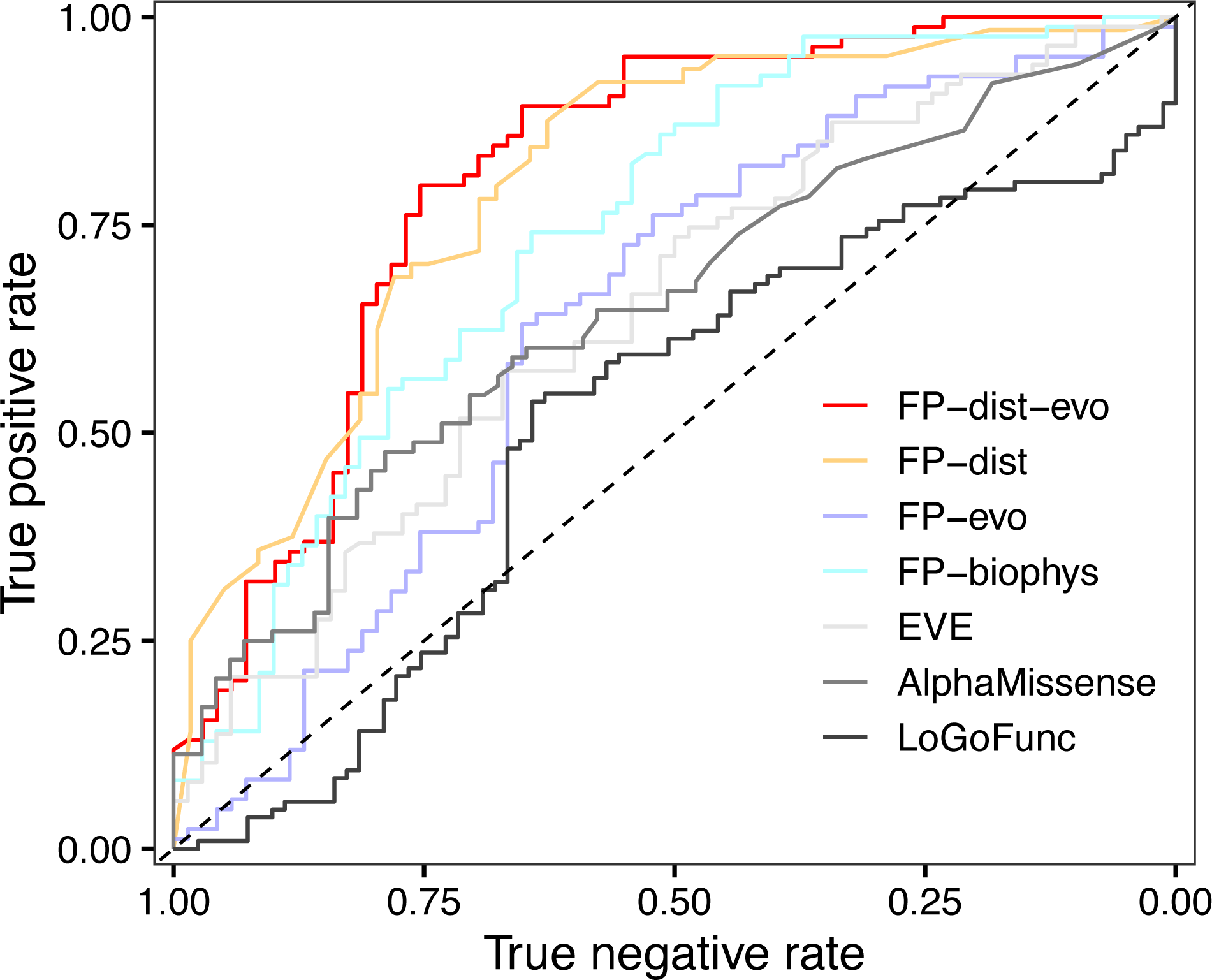
ROC curve for the four ML-based functional predictors (PP-dist, PP-evo, PP-biophys, PP-dist&evo), the EVE score and AlphaMissense pathogenicity predictor, and the LoGoFunc LoF/GoF predictor on the dataset composed of 159 variants in the GRIN genes.

If we combine distance and evolutionary scores, our final best functional predictor for GRIN variants, FP-dist&evo reaches an overall accuracy of 0.765 and a MCC of 0.523 (for predictions see Supplementary Table 1). As a comparison, are also reported in Table 4 the performances of the pathogenicity predictor Alphamissense and of the only other available non-protein specific functional (LoF/GoF) predictor LoGoFunc (https://www.biorxiv.org/content/10.1101/2022.06.08.495288v1.full.pdf). These performances show that pathogenicity prediction scores like Alphamissense have little to no predictive power for the functional effects of variants and FP-dist&evo clearly outperforms, on the GRIN variants, the only other available functional predictor.

### Global incidence estimates of increase/decrease subdisorders

We previously published a mutation-rate-informed prediction model to predict the birth incidence of *de novo* variant-associated disorders such as GRIN disorders. However, we were not able to separate GoF vs LoF disorder subtypes. In the final set of analyses, we estimated the fraction of molecular mechanism defined disorder subtypes using two approaches. In the first, we predicted for all observed variants in our expert curated patient cohort the “increasing” and “decreasing” function (Supplementary Table 1) status (75% most confident class assignments). The fraction for either classification was used as factor for the previous gene level birth incidences. Among our patient cohort, using our increased/decreased functional predictor, we predicted an increased functional effect in 52% of all cases in *GRIN1*. In contrast, in patient variants observed in *GRIN2A* and *GRIN2B* the more recurrent predicted functional effect was “decreasing” function (Decreasing *_GRIN2A_*= 61%, Decreasing *_GRIN2B_*= 56%, Figure 7B). Since the first approach might be confounded by a not yet recognized ascertainment bias of our cohort, we used a second approach to estimate the ratio of variants that “increase”/”decrease” function in the patient population. We applied the functional prediction algorithm to all possible variants along *GRIN1*, *GRIN2A* and *GRIN2B* which we classified as pathogenic using our pathogenicity prediction algorithm (N = 4,630). “Decreasing” effects are suggested as the dominant functional effect in each of the three GRIN genes (Figure 7C). Notably, this model doesn’t consider that some variants might occur de novo more frequent that others due to CpG rich codons. Therefore, our two approaches, that each have advantages and disadvantages, we estimate the incidence for “increasing” function variant associated disorders for *GRIN1*as 2.15-2.81, for *GRIN2A* as 0.32-1.08 and for *GRIN2B* 0.50-2.31 in 100,000 births. Similarly, for “Decreasing” variant associated disorders we obtained an incidence estimate for *GRIN1* of 2.63-3.33, for *GRIN2A* as 1.7-2.46 and for *GRIN2B* 3.03-4.85 in 100,000 births. Inclusion of measures of surface localization together with parameters used to define “Increasing” and “Decreasing” function are needed before LoF and GoF can formally be used.

**Figure 7.**
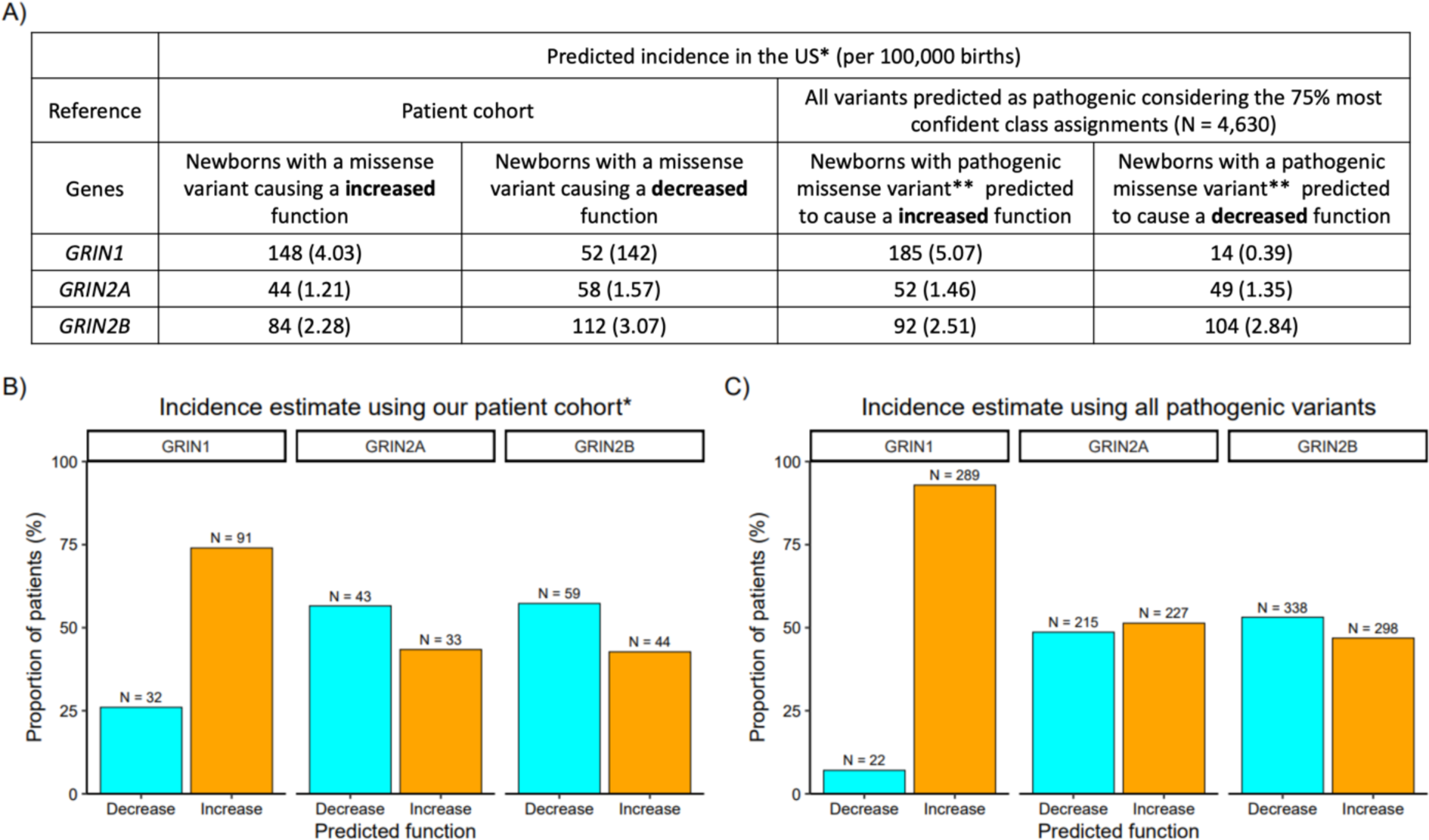
Prediction of the missense variant effects across our cohort of patient variants. **A)** Incidence estimates for the functional effect of NMDAR encoding genes. Overall incidences were obtained from (López-Rivera *et al., 2020*)^83^. Function specific incidences were calculated based on the gene-wise observed ratio of variants with an Increasing/Decreasing effect. *according to 3,659,289 births in USA. ** predicted with PP-dist&evo pathogenicity model. **B)** Gene wise fraction of patient variants from our patient cohort (see methods for details) predicted to cause an increasing or decreasing function considering the 75% most confident class assignments. **C)** As in **B** but predicting the function of all variants in *GRIN1*, *GRIN2A* and *GRIN2B* that were predicted to be pathogenic using our best pathogenicity prediction method, PP-dist&evo (considering the 75% most confident class assignments).

## Discussion

Tailoring treatment to individual patients’ genetic variants has made significant progress in many fields of medicine in recent years ^84^. For disorders caused by genetic variants in GRIN genes, the possibility of successful drug treatment critically depends on the knowledge of the change in function caused by the pathogenic variant, that is gain or loss (partial or complete) of the NMDAR function. Indeed, the knowledge of the molecular mechanism affected by a variant can guide safe and effective targeted treatment^45^. To date, only 20% of all the single amino acid exchanges in GRIN genes have been characterized by electrophysiological and biochemical readouts^85^. In this work we collected the most comprehensive dataset of GRIN variants located in the structurally resolved regions of the NMDAR, with 201 patient variants curated by clinical and genetics experts together and 631 population variants and 159 variants whose molecular consequences have been characterized by in vitro electrophysiology. Taking advantage of this comprehensive clinical and functional dataset of GRIN variants and using machine learning, we identify distance from ligands as a main predictive feature for both pathogenicity and functional prediction and we provide the most accurate pathogenicity prediction specifically developed for GRIN variants and the first computational method for the prediction of variant functional consequences in the GRIN genes (for prediction scores see Supplementary Table 1).

On the one hand, protein-unspecific pathogenicity predictors take the advantage of large training datasets and of capturing general principles of protein functioning though the use of massive evolutionary information^78,86–88^. On the other hand, protein-specific pathogenicity predictors that can incorporate knowledge on protein-specific structure and function, have been shown to enhance the accuracy of pathogenicity prediction^50,51,89,90^. When allowed by data availability, protein-specific pathogenicity predictors can improve the accuracy of pathogenicity prediction, by capturing protein-specific structural patterns and constraints. Here we increase the accuracy of pathogenicity prediction for missense variants in the GRIN genes by integrating structural information on the distance of variant residues to functionally important sites, ligands and the pore.

We also showed that distance from ligands and the pore has predictive power also for functional prediction. A limitation of this work, however, is that we explicitly omit effects of variants on trafficking, which almost certainly have structural determinants beyond the ion channel pore and agonist binding pocket. Surface expression is clinically important, and a topic we will explore separately. So, our functional predictor is predicting increased and decreased function and not GoF/LoF. In line with previously reported observations in which variants with a LoF effect are predominantly located in the ABD domain, due to a disturbance of the agonist binding sites of GluN1 and GluN2A/B^4,14^ we also find an enrichment of variants with increased functional effect. For a couple of variants close to the ligand binding site it has been proposed that the amino acid substitutions lead to a reduced agonist binding^59,60^. In contrast most variants in the TMD have been shown to have a GoF (or complex) effect^91–93^. Here we quantify for the first time the correlation of the functional effects of variants in NMDAR proteins with their spatial distance from the ligands. Hence these 3D distances can be used as a proxy to estimate the functional consequence of yet untested variants. Although this is the first step towards an accurate model specifically designed to predict the functional effect of NMDAR variants, we show that protein-unspecific models trained for pathogenicity were not sufficient to develop a strong prediction model for variant effects in NMDAR genes, and only including our newly generated distance features sufficiently boosts the performances to allow functional prediction.

This work is affected by the limitations that classification of the functional consequence of a variant in the two classes of LoF and GoF is an over-simplification of the real biophysical modifications which take place on a molecular level. Still, we could separately show that the distance of variant residues to ligands that regulate the NMDAR in particular correlate with the ligand-specific fold change potency, demonstrating its validity also on the level of individual electrophysiological readouts. Consequently, once more specific electrophysiological readouts will become available our spatial distance to functional site annotation will become even more helpful to train models that predict the molecular consequence with higher granularity.

In summary, we introduced a potentially powerful approach to predict the directionality of the functional effects of likely pathogenic missense variants in GRIN genes. In a clinical setting like, treatment decisions must often be made before functional studies of disease-causing variants can be done. In the future, our prediction method could be adapted and benchmarked for use in conjunction with best current clinical practices, for example, to predict which individuals with pathogenic variants may be likely to benefit from a particular treatment based on their variants’ LoF or GoF effects. Our method could potentially be refined with large-scale experimental data, for example, by introducing more specific types of predictions than the mere binary LoF and GoF classification, such as directly predicting a specific change of potency (e.g., glutamate). Because most GRIN genes are depleted for functional variants in the general population, it is likely that more GRIN genes could contribute to disease for which disease associations or mechanisms have not yet been elucidated and to which our method could potentially be applied – such as LoF variants in *GRIN2D*. In future iterations, also clinical and phenotypic data might be incorporated to enhance predictions for the underlying molecular defect.

## Data Availability

All data produced in the present work are contained in the manuscript

## Acknowledgements

We thank Rui Song, Wenshu XiangWei, Yuchen Xu, Daniel Teuscher, Lingling Xie, Ruth K. Mizu, and Wenjuan Chen for valuable assistance and sharing unpublished data.

## Disclosures

H.Y. and S.F.T. are co-inventors of Emory-owned intellectual property. S.F.T. is a member of the SAB for Sage Therapeutics, Eumentis Therapeutics, Neurocrine, the GRIN2B Foundation, the CureGRIN Foundation, and CombinedBrain. S.F.T. is a consultant for GRIN Therapeutics. H.Y. is the PI on a research grant from Sage Therapeutics and GRIN Therapeutics to Emory. S.F.T. is PI on a research grant from GRIN Therapeutics to Emory. S.F.T. is cofounder of NeurOp, Inc. and Agrithera. DL recieved funds from the Simons Foundation and GRIN Therapeutics.

## Grant Support

This work was supported by NS111619 (ST), HD082373 (HY), and AG072142 (SJM).

## Notes

### Summary of Updates

The supplementary data has been updated and typos in the author's names was corrected.

