## Supplemental Table S2 for "Distances from ligands as main predictive features for pathogenicity and functional effect of variants in NMDA receptors"

**Supplemental Table S2: Missense variants for which analysis needed for GoF or LoF call described by Myers et al. (2023) is complete**

| Gene | Residue Position | WT Amino Acid | Variant Amino Acid | Call for this paper | Formal Call by Myers et al criteria | ClinVar | PMID | Reference |
|---|---|---|---|---|---|---|---|---|
| *GRIN1* | 531 | Lys | Arg | Decrease | Likely LoF* | VCV002686033.1 | -- | this study |
| *GRIN1* | 532 | Pro | His | Decrease | Likely LoF | VCV000421469 | 34413877, 37369021 | Zhang 2021, Myers 2023 |
| *GRIN1* | 548 | Arg | Gln | Increase | Likely GoF | VCV000521298 | 37000222, 37369021 | Xie 2023, Myers 2023 |
| *GRIN1* | 549 | Ser | Arg | Decrease | Likely LoF | VCV000520575 | 37000222, 37369021 | Xie 2023, Myers 2023 |
| *GRIN1* | 551 | Leu | Pro | Increase | Likely GoF | -- | 29365063, 37000222, 37369021 | Fry 2018; Xie 2023, Myers 2023 |
| *GRIN1* | 556 | Gln | Arg | Increase | Likely GoF | VCV000976358 | -- | this study |
| *GRIN1* | 557 | Pro | Leu | Decrease | Likely LoF* | VCV000372881 | 37000222 | Xie 2023 |
| *GRIN1* | 557 | Pro | Arg | Increase | Possible GoF | VCV000235846 | 28095420 | this study, Ogden 2017 |
| *GRIN1* | 559 | Gln | Arg | Increase | Possible GoF | -- | 37000222, 37369021 | Xie 2023, Myers 2023 |
| *GRIN1* | 637 | Ala | Ser | Increase | Likely GoF | -- | 38538865 | Xu 2023 |
| *GRIN1* | 637 | Ala | Val | Increase | Possible GoF | VCV000452518 | 38538865 | Xu 2023 |
| *GRIN1* | 638 | Gly | Ala | Increase | Likely GoF | VCV000975942.5 | 38538865 | Xu 2023 |
| *GRIN1* | 638 | Gly | Val | Increase | Possible GoF | -- | 38538865 | Xu 2023 |
| *GRIN1* | 638 | Gly | Ser | Increase | Possible GoF | VCV001502674 | -- | this study |
| *GRIN1* | 641 | Met | Ile | Increase | Possible GoF | VCV000403957 | 25864721, 30355546, 37369021, 38538865 | Ohba 2015, Pironti 2018, Myers, Xu 2023 |
| *GRIN1* | 641 | Met | Leu | Increase | Likely GoF | VCV001700197 | 30355546, 38538865 | Pironti 2018, Xu 2023 |
| *GRIN1* | 642 | Ile | Leu | Decrease | Likely LoF | VCV000421935 | 38538865 | Xu 2023 |
| *GRIN1* | 642 | Ile | Thr | Decrease | Possible LoF | -- | 38538865 | Xu 2023 |
| *GRIN1* | 643 | Ala | Val | Increase | Likely GoF | VCV000539837.6 | 38538865 | Xu 2023 |
| *GRIN1* | 644 | Val | Met | Increase | Likely GoF | VCV000451906 | 38538865 | Xu 2023 |
| *GRIN1* | 645 | Ala | Ser | Increase | Possible GoF | VCV000917873 | 27164704, 38538865 | Lemke 2016, Xu 2023 |
| *GRIN1* | 650 | Asn | Ile | Increase | Possible GoF | VCV000429784 | 38538865 | Xu 2023 |
| *GRIN1* | 650 | Asn | Lys | Increase | Possible LoF | VCV000487502; VCV | 25864721, 38538865 | Ohba 2015, Xu 2023 |
| *GRIN1* | 652 | Ala | Thr | Decrease | Likely LoF | -- | 38538865 | Xu 2023 |
| *GRIN1* | 653 | Ala | Gly | Increase | Indeterminant | VCV000521282 | 27164704, 38538865 | Lemke 2016, Xu 2023 |
| *GRIN1* | 653 | Ala | Thr | Increase | Likely GoF | -- | 35393335, 38538865 | Brock 2023, Xu 2023 |
| *GRIN1* | 654 | Phe | Cys | Increase | Likely GoF | VCV000559643 | 38538865 | Xu 2023 |
| *GRIN1* | 655 | Leu | Gln | Increase | Likely GoF | VCV000561020 | 38538865 | Xu 2023 |
| *GRIN1* | 744 | Cys | Gly | Increase | Likely GoF | -- | -- | this study |
| *GRIN1* | 806 | Ala | Glu | Increase | Likely GoF | VCV000522682 | -- | this study |
| *GRIN1* | 844 | Arg | Leu | Increase | Possible GoF | VCV000598966 | -- | this study |
| *GRIN2A* | 483 | Gly | Arg | Decrease | Likely LoF | VCV000812185 | 27839871, 28242877, 37369021 | Swanger 2016, Addis 2017, Myers 2023 |
| *GRIN2A* | 506 | Val | Ala | Increase | Possible GoF | VCV000205650 | 27839871, 37369021 | Swanger 2016, Myers 2023 |
| *GRIN2A* | 511 | Ser | Leu | Decrease | Likely LoF* | VCV000080290 | -- | this study |
| *GRIN2A* | 518 | Arg | His | Decrease | Likely LoF* | VCV000088732 | 27839871, 37369021 | Swanger 2016, Myers 2023 |
| *GRIN2A* | 518 | Arg | Cys | Decrease | Likely LoF* | VCV000205687 | -- | this study |
| *GRIN2A* | 531 | Thr | Met | Decrease | Likely LoF* | VCV000088730 | 27839871, 37369021 | Swanger 2016, Myers 2023 |
| *GRIN2A* | 545 | Ser | Leu | Decrease | Possible LoF | -- | 37000222, 37369021 | Xie 2023, Myers 2023 |
| *GRIN2A* | 548 | Ala | Pro | Decrease | Likely LoF* | VCV000390621 | 37000222, 37369021 | Xie 2023, Myers 2023 |
| *GRIN2A* | 551 | Glu | Lys | Increase | Likely GoF | VCV000520918 | 37000222, 37369021 | Xie 2023, Myers 2023 |
| *GRIN2A* | 552 | Pro | Arg | Increase | Possible GoF | VCV000039663 | 28095420, 37369021 | Ogden 2017, Myers 2023 |
| *GRIN2A* | 554 | Ser | Thr | Increase | Likely GoF | VCV001708207 | 37000222, 37369021 | Xie 2023, Myers 2023 |
| *GRIN2A* | 611 | Leu | Gln | Increase | Likely GoF | VCV001325863 | 31429998, 37369021 | Li 2019, Myers 2023 |
| *GRIN2A* | 615 | Asn | Lys | Increase | Possible GoF | VCV000029733 | 20890276, 31429998, 37369021 | Endele 2010, Li 2019, Myers 2023 |
| *GRIN2A* | 635 | Ala | Thr | Increase | Likely GoF | VCV000985807.3 | 38538865 | Xu 2023 |
| *GRIN2A* | 639 | Val | Ile | Increase | Likely GoF | VCV001498958 | 38538865 | Xu 2023 |
| *GRIN2A* | 642 | Leu | Met | Increase | Possible GoF | VCV002499601.2 | 38538865 | Xu 2023 |
| *GRIN2A* | 642 | Leu | Arg | Increase | Possible GoF | VCV000985631 | 25590979, 38538865 | Zhu 2015, Xu 2023 |
| *GRIN2A* | 643 | Ala | Asp | Increase | Possible GoF | VCV000243097 | 29644724, 38538865 | Fernandez-Marmiesse 2018, Xu 2023 |
| *GRIN2A* | 644 | Ser | Gly | Increase | Possible GoF | VCV001325864 | 32577763, 38538865 | Amador 2020, Xu 2023 |

| | | | | | | | | |
|---|---|---|---|---|---|---|---|---|
| *GRIN2A* | 646 | Thr | Ala | Increase | Possible GoF | VCV000488524 | 28109652, 30544257, 38538865 | von Stulpnagel 2017, Strehlow 2019, Xu 2023 |
| *GRIN2A* | 646 | Thr | Arg | Increase | Likely GoF | VCV000432031 | 38538865 | Xu 2023 |
| *GRIN2A* | 648 | Asn | Ser | Decrease | Possible LoF | VCV000205652 | 26633542, 30544257, 38538865 | Retterer 2016, Strehlow 2019, Xu 2023 |
| *GRIN2A* | 649 | Leu | Val | Increase | Possible GoF | VCV000039662 | 23033978, 30544257, 38538865 | de Ligt 2012, Strehlow 2019, Xu 2023 |
| *GRIN2A* | 650 | Ala | Ser | Increase | Possible GoF | VCV001021787.9 | 38538865 | Xu 2023 |
| *GRIN2A* | 652 | Phe | Val | Decrease | Possible LoF | VCV000088733 | 23933820, 38538865 | Lesca 2013, Xu 2023 |
| *GRIN2A* | 653 | Met | Ile | Increase | Indeterminant | VCV001325867 | 30544257, 38538865 | Strehlow 2019, Xu 2023 |
| *GRIN2A* | 653 | Met | Val | Increase | Possible GoF | VCV001325866 | 30544257, 38538865 | Strehlow 2019, Xu 2023 |
| *GRIN2A* | 654 | Ile | Thr | Increase | Possible GoF | VCV000941334 | 30544257, 38538865 | Strehlow 2019, Xu 2023 |
| *GRIN2A* | 690 | Thr | Met | Decrease | Likely LoF | VCV000444362 | -- | this study |
| *GRIN2A* | 694 | Ile | Thr | Decrease | Likely LoF | VCV001325869 | 27839871 | this study, Swanger 2016 |
| *GRIN2A* | 695 | Arg | Gln | Increase | Likely GoF | VCV000205689 | -- | this study |
| *GRIN2A* | 705 | Met | Val | Decrease | Possible LoF | VCV001325870 | 27839871, 37369021 | Swanger 2016, Myers 2023 |
| *GRIN2A* | 716 | Ala | Thr | Decrease | Likely LoF | VCV000205656 | 27839871, 37369021 | Swanger 2016, Myers 2023 |
| *GRIN2A* | 716 | Ala | Asp | Decrease | Likely LoF* | VCV000375534 | 23933820 | this study, Lesca 2013, |
| *GRIN2A* | 731 | Asp | Asn | Decrease | Likely LoF | VCV000205657 | 27839871, 28242877, 28182669, 37369021 | Swanger 2016, Addis 2017, Gao 2017, Myers 2023 |
| *GRIN2A* | 734 | Val | Leu | Decrease | Possible LoF | VCV000433123 | 27839871, 37369021 | Swanger 2016, Myers 2023 |
| *GRIN2A* | 772 | Lys | Glu | Decrease | Likely LoF | VCV001483207 | 23933819, 27839871 | this study, Lemke 2013, Swanger 2016 |
| *GRIN2A* | 812 | Leu | Met | Increase | Likely GoF | VCV001325703 | 24504326 | this study, Yuan 2014 |
| *GRIN2A* | 817 | Met | Val | Increase | -- | VCV000205659.4 | 28126851, 30544257 | Chen 2017, Strehlow 2019 |
| *GRIN2A* | 818 | Ala | Val | Increase | Likely GoF | VCV001065504 | -- | this study |
| *GRIN2B* | 413 | Glu | Gly | Decrease | Likely LoF | VCV000143189 | 24863970, 27839871, 37369021 | Adams 2014, Swanger 2016, Well 2018, Myers 2023 |
| *GRIN2B* | 461 | Cys | Phe | Decrease | Possible LoF | VCV000916598 | 27839871, 29511171, 37369021 | Swanger 2016, Fedele 2018, Myers 2023 |
| *GRIN2B* | 533 | Gly | Asp | Decrease | Likely LoF | VCV000397525 | 37369021 | Myers 2023 |
| *GRIN2B* | 540 | Arg | His | Increase | Indeterminant | VCV000162087 | 27839871, 37369021 | Swanger 2016, Myers 2023 |
| *GRIN2B* | 541 | Ser | Gly | Increase | Possible GoF | VCV001708206 | 37000222, 37369021 | Xie 2023, Myers 2023 |
| *GRIN2B* | 541 | Ser | Arg | Decrease | Likely LoF | VCV000916601 | 28377535, 37000222, 37369021 | Platzer 2017, Xie 2023, Myers 2023 |
| *GRIN2B* | 543 | Gly | Arg | Increase | Possible GoF | VCV000984886 | 37000222 | Xie 2023 |
| *GRIN2B* | 549 | Ala | Val | Decrease | Likely LoF* | VCV001723180 | 37000222, 37369021 | Xie 2023, Myers 2023 |
| *GRIN2B* | 550 | Phe | Ser | Decrease | Likely LoF* | VCV000522093 | 37000222, 37369021 | Xie 2023, Myers 2023 |
| *GRIN2B* | 551 | Leu | Ser | Decrease | Likely LoF* | | 37000222, 37369021 | Xie 2023, Myers 2023 |
| *GRIN2B* | 553 | Pro | Thr | Decrease | Likely LoF | | 37000222, 37369021 | Xie 2023, Myers 2023 |
| *GRIN2B* | 553 | Pro | Leu | Decrease | Likely LoF* | VCV000039661 | 28095420, 29511171, 37369021 | Ogden 2017, Fedele 2018, Myers 2023 |
| *GRIN2B* | 555 | Ser | Ile | Decrease | Likely LoF* | VCV000916602 | 37000222, 37369021 | Xie 2023, Myers 2023 |
| *GRIN2B* | 555 | Ser | Asn | Increase | Likely GoF | VCV001483642 | 37000222, 37369021 | Xie 2023, Myers 2023 |
| *GRIN2B* | 558 | Val | Ile | Decrease | Likely LoF | VCV000374243.11 | 28377535, 29681796 | Platzer 2017; Vyklicky 2018 |
| *GRIN2B* | 559 | Trp | Arg | Decrease | Likely LoF* | VCV000984850 | -- | this study |
| *GRIN2B* | 559 | Trp | Cys | Decrease | Likely LoF* | -- | -- | this study |
| *GRIN2B* | 607 | Trp | Cys | Increase | Possible GoF | VCV000916606 | 29681796, 31429998, 37369021 | Vyklicky 2018, Li 2019, Myers 2023 |
| *GRIN2B* | 607 | Trp | Ser | Increase | Possible GoF | VCV000828156 | 37369021 | Myers 2023 |
| *GRIN2B* | 611 | Gly | Val | Increase | Likely GoF | VCV000205730 | 28377535, 31429998, 37369021 | Platzer 2017, Li 2019, Myers 2023 |
| *GRIN2B* | 615 | Asn | Lys | Increase | Possible GoF | VCV000916607 | 28377535, 31429998, 37369021 | Platzer 2017, Li 2019, Myers 2023 |
| *GRIN2B* | 615 | Asn | Ile | Increase | Possible GoF | VCV000162086 | 29511171, 29681796, 31429998, 37369021 | Fedele 2018, Vyklicky 2018, Li 2019, Myers 2023 |
| *GRIN2B* | 616 | Asn | Lys | Increase | Possible GoF | VCV000916608 | 28377535, 31429998, 37369021 | Platzer 2017, Li 2019, Myers 2023 |
| *GRIN2B* | 616 | Asn | Ile | Increase | Likely GoF | -- | -- | this study |
| *GRIN2B* | 616 | Asn | Ser | Increase | Possible GoF | VCV000843669 | -- | this study |
| *GRIN2B* | 620 | Val | Met | Increase | Possible GoF | VCV000205710 | 31429998, 37369021 | Li 2019, Myers 2023 |
| *GRIN2B* | 639 | Ala | Val | Increase | Possible GoF | VCV000208749 | 25356970, 26795593, 28377535, 38538865 | Farwell 2015, Helbig 2016, Platzer 2017, Xu 2023 |
| *GRIN2B* | 641 | Ile | Thr | Increase | Indeterminant | VCV000985189 | 38538865 | Xu 2023 |
| *GRIN2B* | 652 | Ala | Gly | Increase | Likely GoF | VCV000245714 | 38538865 | Xu 2023 |
| *GRIN2B* | 652 | Ala | Pro | Decrease | Likely LoF* | VCV000452873.2 | 38538865 | Xu 2023 |
| *GRIN2B* | 654 | Met | Ile | Increase | Possible GoF | -- | -- | this study |
| *GRIN2B* | 654 | Met | Val | Increase | Possible GoF | -- | -- | this study |
| *GRIN2B* | 655 | Ile | Phe | Increase | Indeterminant | VCV000916612 | 28377535, 38538865 | Platzer 2017, Xu 2023 |
| *GRIN2B* | 682 | Arg | His | Increase | Possible GoF | VCV000268209 | 37369021 | Myers 2023 |
| *GRIN2B* | 689 | Gly | Cys | Decrease | Likely LoF* | -- | 34212862 | Kellner 2021 |

| | | | | | | | | |
|---|---|---|---|---|---|---|---|---|
| ***GRIN2B*** | 689 | Gly | Ser | **Decrease** | **Likely LoF** | VCV000224818 | 34212862, 37369021 | Kellner 2021, Myers 2023 |
| ***GRIN2B*** | 691 | Thr | Ala | **Decrease** | **Likely LoF*** | VCV000561023 | -- | this study |
| ***GRIN2B*** | 691 | Thr | Ile | **Decrease** | **Likely LoF*** | -- | -- | this study |
| ***GRIN2B*** | 693 | Arg | Ser | **Increase** | **Likely GoF** | VCV000916586 | 37369021 | Myers 2023 |
| ***GRIN2B*** | 695 | Ile | Thr | **Decrease** | **Likely LOF** | VCV000234790 | 37369021 | Myers 2023 |
| ***GRIN2B*** | 695 | Ile | Ser | **Decrease** | **Likely LOF** | VCV000432191 | 37369021 | Myers 2023 |
| ***GRIN2B*** | 696 | Arg | His | **Increase** | **Likely GoF** | VCV000489393 | 27839871, 37369021 | Swanger 2016, Myers 2023 |
| ***GRIN2B*** | 696 | Arg | Cys | **Increase** | **Likely GoF** | VCV002574431.1 | -- | this study |
| ***GRIN2B*** | 706 | Met | Val | **Decrease** | **Possible LOF** | VCV000374226 | 37369021 | Myers 2023 |
| ***GRIN2B*** | 717 | Ala | Glu | **Decrease** | **Likely LoF*** | -- | -- | this study |
| ***GRIN2B*** | 746 | Cys | Trp | **Decrease** | **Likely LoF** | VCV001694662 | -- | this study |
| ***GRIN2B*** | 797 | Leu | Phe | **Decrease** | **Likely LoF** | -- | -- | this study |
| ***GRIN2B*** | 807 | Glu | Lys | **Decrease** | **Likely LoF** | VCV000916587 | 37369021 | Myers 2023 |
| ***GRIN2B*** | 810 | Ser | Asn | **Increase** | **Likely GoF** | VCV000800894 | 37369021 | Myers 2023 |
| ***GRIN2B*** | 813 | Leu | Val | **Increase** | **Likely GoF** | VCV000864861 | 35393335 | this study, Brock 2023 |
| ***GRIN2B*** | 818 | Met | Arg | **Decrease** | **Likely LoF*** | VCV000411110 | -- | this study |
| ***GRIN2B*** | 818 | Met | Thr | **Increase** | **Possible GoF** | VCV000245960 | 28377535, 35393335 | this study, Platzer 2017, Brock 2023 |
| ***GRIN2B*** | 820 | Gly | Ala | **Decrease** | **Likely LOF** | VCV000208643.19 | 34008892, 30217972, 28377535 | Marinakis 2021, Amin 2018, Platzer 2017 |
| ***GRIN2B*** | 820 | Gly | Glu | **Decrease** | **Likely LoF*** | VCV000580700 | 28377535, 29681796, 30217972 | Platzer 2017, Vyklicky 2018, Amin 2018 |
| ***GRIN2B*** | 821 | Val | Phe | **Decrease** | **Possible LoF** | VCV000984853 | 37369021 | Myers 2023 |
| ***GRIN2B*** | 824 | Met | Arg | **Decrease** | **Likely LoF*** | VCV000561027 | 28377535, 29681796 | Platzer 2017, Vyklicky 2018 |
