## Supplemental Table S5 for "Distances from ligands as main predictive features for pathogenicity and functional effect of variants in NMDA receptors"

**Supplemental Table S5: Data describing missense variant effects on NMDAR properties.**

| *GRIN* -variant | Glutamate $EC_{50}$ (95% CI), μM | Glycine $EC_{50}$ (95% CI), μM | $Mg^{2+}$ $IC_{50}$ (95% CI), μM | $P_{OPEN}$ | $\tau_{WEIGHT}$ (ms) | Surface Expression (% of total) |
|---|---|---|---|---|---|---|
| **WT 1/2A*** | **3.6** | **1.2** | **20** | **0.23** | **60** | **100** |
| **1-K531R/2A** | 9.9 (8.9, 11.1) | 2.7 (2.4,2.9) | 18 (14, 24) | 0.098 ± 0.0052 | 26 ± 4.1 | 113 ± 12 |
| **1-Q556R/2A** | 1.1 (0.64, 1.9) | 0.42 (0.34, 0.51) | 29 (22, 39) | 0.48 ± 0.018 | 200 ± 49 | 113 ± 3.1 |
| **1-P557R/2A[a]** | 0.37 (0.34, 0.41) | 0.088 (0.080, 0.10) | 28 (20, 39) | 0.026 ± 0.0016 | 395 ± 32 | 39 ± 0.099 |
| **1-G638S/2A** | 3.3 (2.6, 4.1) | 1.1 (0.95, 1.3) | 83 (58, 119) | 0.24 ± 0.010 | 54 ± 5.6 | 60 ± 16 |
| **1-C744G/2A** | 0.71 (0.51, 1.0) | 1.3 (1.1, 1.5) | 26 (21, 30) | 0.33 ± 0.011 | 163 ± 16 | 101 ± 6.7 |
| **1-A806E/2A** | 0.28 (0.23, 0.33) | 0.073 (0.055, 0.10) | 18 (16, 21) | 0.30 ± 0.011 | 441 ± 64 | 116 ± 1.6 |
| **1-R844L/2A** | 2.4 (2.1, 2.9) | 0.88 (0.76, 1.0) | 54 (46, 63) | 0.35 ± 0.022 | 98 ± 13 | 140 ± 10 |
| **WT 1/2A*** | **3.2** | **1.2** | **26** | **0.17** | **54** | **100** |
| **2A-R518C** | 3600 (3260, 4000) | 1.5 (1.2, 1.9) | 21 (17, 25) | tstm | tstm | 32 ± 4.0 |
| **2A-T690M** | 5900 (4700,7300) | 1.4 (1.1, 1.7) | 29 (24,34) | tstm | tstm | 30 ± 5.2 |
| **2A-I694T[b]** | 9.7 (9.0, 11) | 0.87 (0.63, 1.2) | 28 (20,40) | 0.13 ± 0.0060 | 45 ± 3.7 | 72 ± 8.0 |
| **2A-R695Q** | 0.32 (0.28, 0.35) | 1.2 (1.1, 1.4) | 15 (11, 21) | 0.20 ± 0.01 | 257 ± 34 | 77 ± 23 |
| **2A-A716D** | tstm | tstm | tstm | tstm | tstm | n.a. |
| **2A-K772E[b]** | 4.8 (4.4, 5.2) | 1.3 (1.2, 1.5) | 13 (8.5,21) | 0.14 ± 0.005 | 47 ± 5.6 | 18 ± 2.0 |
| **2A-L812M[c]** | 0.40 (0.34, 0.45) | 0.14 (0.12, 0.15) | 36 (32, 41) | 0.40 ± 0.0098 | 443 ± 47 | 85 ± 13 |
| **2A-M817V[d]** | 0.28 (0.20, 0.39) | 0.14 (0.11, 0.17) | 60 (41, 87) | 0.93 ± 0.070 | 632 ± 103 | 100 ± 14 |
| **2A-A818V** | 1.6 (1.3, 2.0) | 0.38 (0.32, 0.45) | 40 (32, 50) | 0.713 ± 0.022 | 86 ± 9.6 | 155 ± 55 |
| **WT 1/2B*** | **1.3** | **0.3** | **23.0** | **0** | **716** | **100** |
| **2B-G543R** | 0.22 (0.19, 0.26) | 0.094 (0.08, 0.12) | 26 (22, 31) | 0.013 ± 0.0023 | 3530 ± 223 | 110 ± 11 |
| **2B-V558I** | 1.0 (0.84, 1.2) | 0.39 (0.33, 0.47) | 24 (20, 30) | 0.0066 ± 0.001 | 376 ± 110 | 103 ± 7.5 |
| **2B-W559C** | tstm | tstm | tstm | tstm | tstm | 74 ± 12 |
| **2B-W559R** | tstm | tstm | tstm | tstm | tstm | 81 ± 1.9 |
| **2B-N616I** | 0.22 (0.14, 0.35) | 0.039 (0.28, 0.55) | >5000 | 0.016 ± 0.0032 | 2240 ± 254 | 82 ± 25 |
| **2B-N616S** | 0.91 (0.81, 1.0) | 0.23 (0.20, 0.28) | >5000 | 0.010 ± 0.00052 | 1140 ± 58 | 83 ± 8.1 |
| **2B-M654I** | 0.059 (0.045, 0.078) | 0.034 (0.022, 0.055) | 42 (30, 59) | 0.032 ± 0.0060 | 5720 ± 647 | 67 ± 11 |
| **2B-M654V** | 0.057 (0.042, 0.077) | 0.087 (0.062, 0.12) | 38 (29, 50) | 0.14 ± 0.020 | 1410 ± 97 | 25 ± 8.4 |
| **2B-T691A** | 1230 (1090, 1380) | 0.46 (0.44, 0.48) | 28 (21, 36) | tstm | tstm | 96 ± 19 |
| **2B-T691I** | 2700 (2490, 2920) | 0.39 (0.32, 0.46) | 22 (16, 29) | tstm | tstm | 119 ± 25 |
| **2B-R696C** | 0.22 (0.19, 0.25) | 0.23 (0.20, 0.28) | 28 (22, 34) | 0.023 ± 0.0020 | 1880 ± 151 | 96 ± 25 |
| **2B-A717E** | tstm | tstm | tstm | tstm | tstm | 41 ± 17 |
| **2B-C746W** | 0.87 (0.72, 1.1) | 2.7 (2.2, 3.3) | 19 (16, 23) | 0.013 ± 0.00089 | 597 ± 55 | 47 ± 8.2 |
| **2B-L797F** | 39 (26, 59) | 0.67 (0.61, 0.74) | 24 (20, 28) | 0.051 ± 0.0031 | 38 ± 4.0 | 89 ± 5.6 |
| **2B-L813V[e]** | 0.27 (0.23, 0.31) | 0.021 (0.013, 0.034) | 24 (21, 29) | 0.021 ± 0.0023 | 1360 ± 101 | 83 ± 7.2 |
| **2B-M818R** | tstm | tstm | tstm | tstm | tstm | 39 ± 8.0 |
| **2B-M818T[e]** | 0.42 (0.30, 0.58) | 0.13 (0.10, 0.19) | 15 (8.5, 26) | 0.15 ± 0.015 | 1680 ± 440 | 63 ± 17 |
| **2B-G820A** | 1.3 (0.96, 1.9) | 0.32 (0.26, 0.39) | 22 (19, 24) | 0.011 ± 0.0016 | 205 ± 10 | 84 ± 7.6 |

* WT values were recorded on the same day for every variant, and the mean for each parameter is given

Mean values for variant receptors are given with 95% confidence intervals determined from the log EC50 or IC50 values, or with SEM. All data are from 8-23 oocytes, 4-15 HEK cells, or 2-10 independent surface expression

tstm indicates the response was too small to measure, n.a. indicates data is not available

a Part of the data set were reanalyzed from data in Ogden et al. 2017 b Part of the data set were reanalyzed from data in Swanger et al. 2016

c Part of the data set were reanalyzed from data in Yuan et al. 2014

d Part of the data set were reanalyzed from data in Chen et al. 2017

e Part of the data set were reanalyzed from data in Brock et al. 2023
