## Supplementary Notes for "Distances from ligands as main predictive features for pathogenicity and functional effect of variants in NMDA receptors"

**ML computation and cross-validation**

The binary classifiers of this study were trained with random forests and implemented through the RandomForestClassifier function of the scikit-learn python package. A combination of 16 estimators, ranging from 10 and 500, and eight different maximum depth parameters, ranging from 2 to 9, were explored. A 5-fold cross-validation was adopted for both the pathogenicity predictor and the Increase/Decrease predictor. Each cross-validation comprises a training, a validation and a test set. The best parameters were chosen as the ones which maximize the Matthews correlation coefficient (MCC) on the validation set. The performances reported in the results section are computed on the test set. Given that distance features are only based on the 3D position of the variant (independently of the substituted residue), two different variants in the same position will have the same distance features. To avoid the risk of overfitting, we pooled variants at the same position (therefore with the same distance features) in the same cross-validation set. For the predictors only based on distance features, only one variant has been retained for each residue position.

**Performance measures for the binary classifiers**

The classifiers were evaluated using the following indexes. For each prediction, the classification for each binary classifier (Pathogenic/Benign and Increase/Decrease) is made at the threshold 0.5. In all the performance measures, TP (true positives) are correctly predicted variants of the first class (Pathogenic or Increase), TN (true negatives) are correctly predicted variants of the second class (Benign or Decrease), FP (false positives) are variants of the second class (Benign or Decrease) that are predicted to belong to the first class (Pathogenic or Increase), and FN (false negatives) are variants of the first class (Pathogenic or Increase) that are predicted to belong to the second class (Benign or Decrease). Predictor performance was evaluated using the following metrics: true positive and negative rates (*TPR, TNR*), positive and negative predicted values (*PPV, NPV*), and overall accuracy (*Q_2_*)

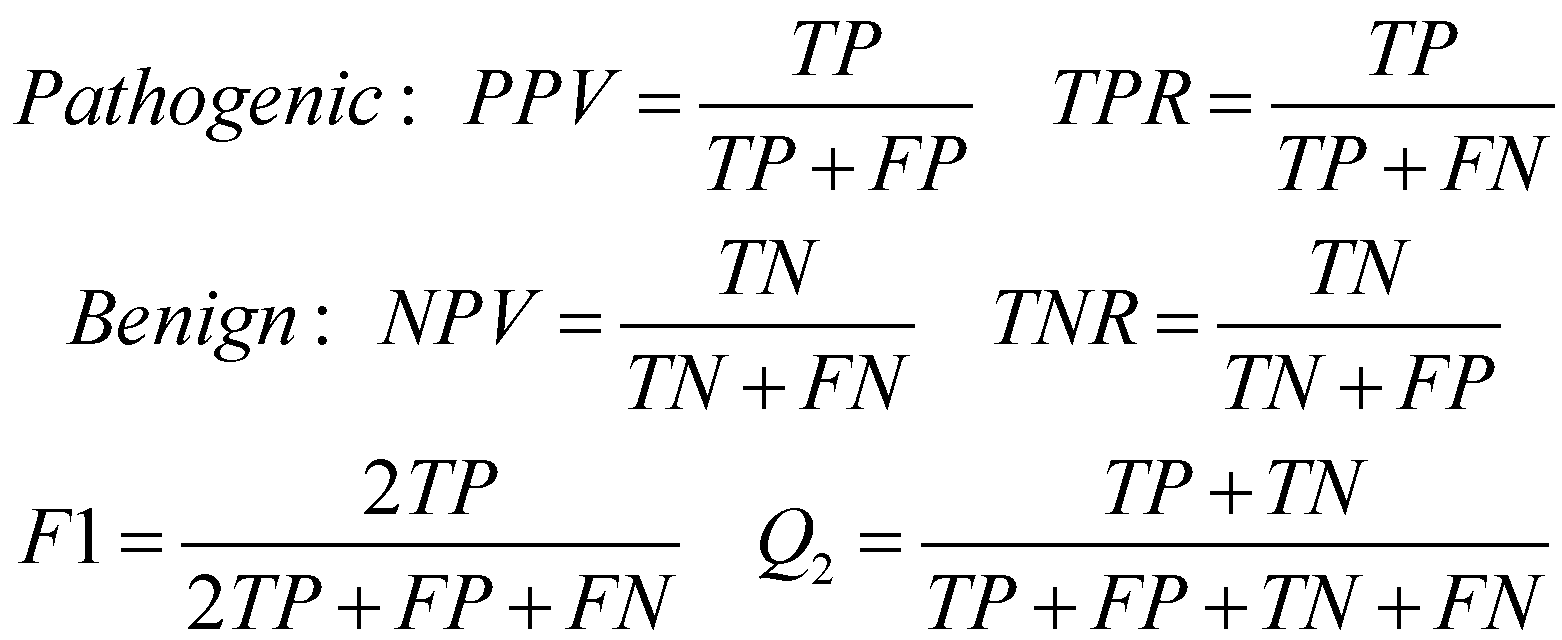

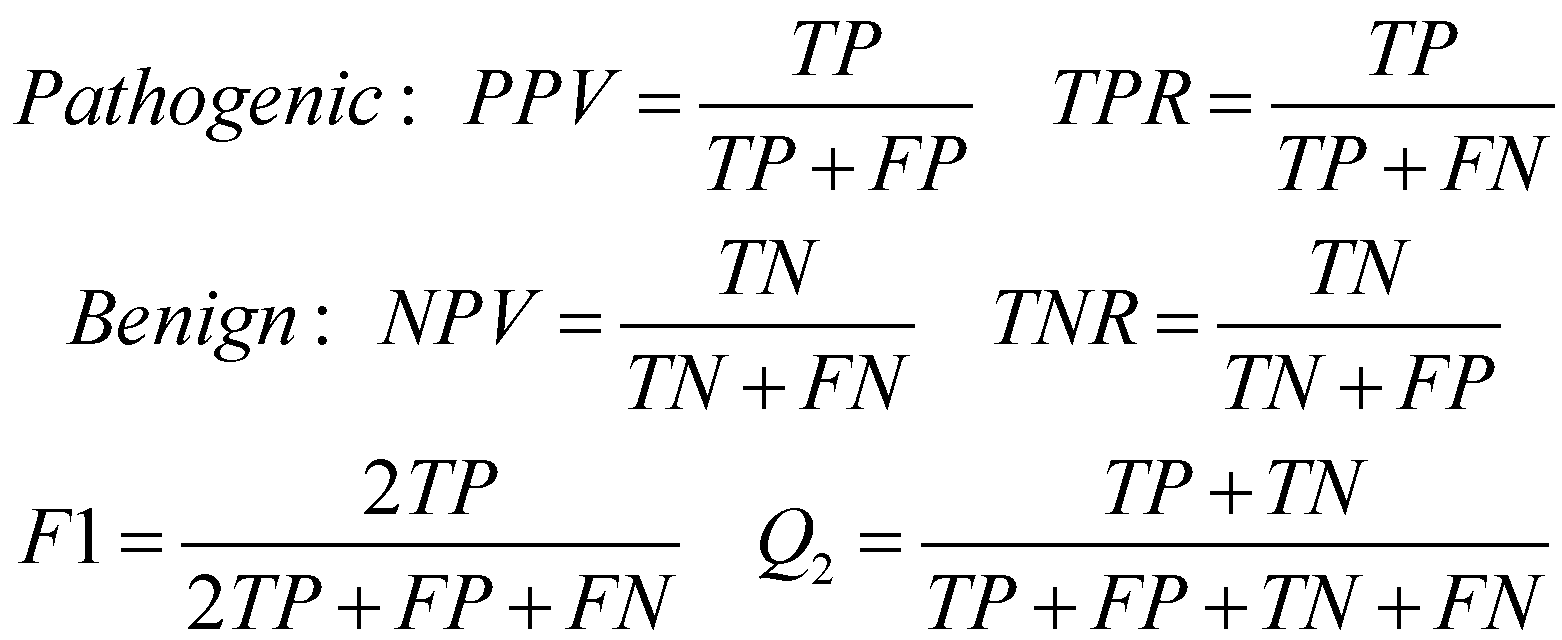

[Eq. 1]

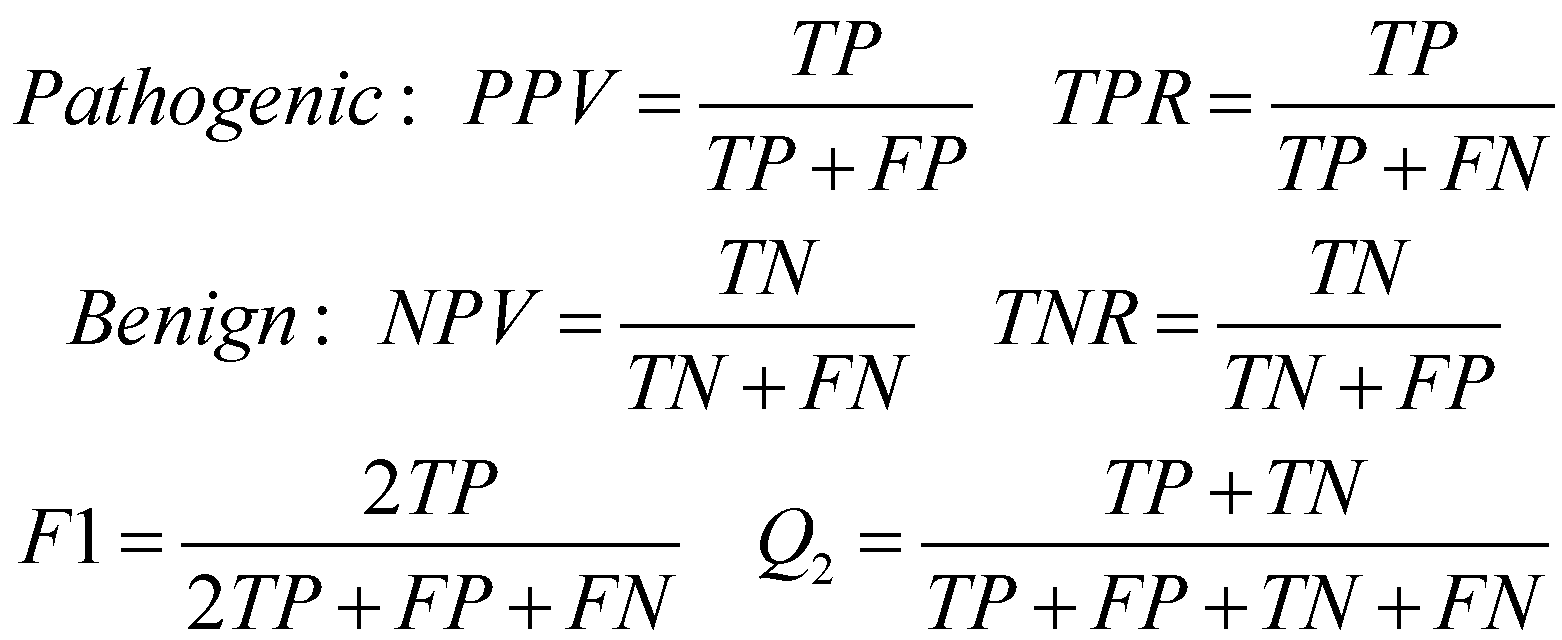

We computed the Matthew’s correlation coefficient MCC (Eq. 2) as:

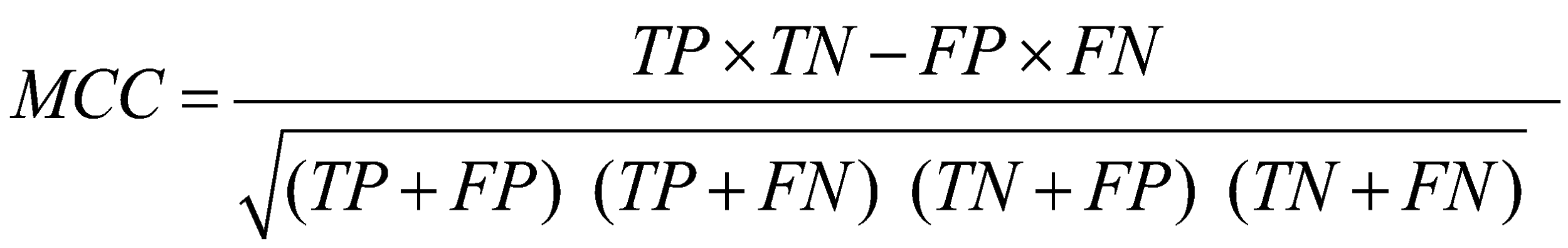

[Eq. 2]

We also calculated the area under the receiver operating characteristic (ROC) curve (AUC), by plotting the True Positive Rate as a function of the False Positive Rate and the Area and the Precision Recall Curve (AUP) at different probability thresholds.
