## Supplemental Table S3 for "Distances from ligands as main predictive features for pathogenicity and functional effect of variants in NMDA receptors"

**Supplemental Table S3. Summary of functional status caused by GRIN variants with partial analysis**

| Gene | Residue Position | WT Amino Acid | Variant Amino Acid | Glutamate Potency | Glycine Potency | $Mg^{2+}$ IC50 | $\tau_{WEIGHT}$ | $P_{OPEN}$ | Classification | ClinVar | PMID | Reference |
|---|---|---|---|---|---|---|---|---|---|---|---|---|
| GRIN1 | 552 | Asp | Glu | H (0.31) | M (0.41) | 1.0 | H (0.23) | H (0.1) | Decrease | VCV000487500.2 | 28095420 | this study, Ogden 2017 |
| GRIN1 | 617 | Ser | Cys | 0.89 | 0.92 | H (5.9) | M (0.43) | H (0.47) | Increase | VCV002579484.1 | 34884460 | this study, Santos-Gómez 2021 |
| GRIN1 | 618 | Gly | Arg | 1.0 | 0.67 | H (>40) | n.a. | H (0.48) | Increase | VCV000981271.3 | 31429998 | Li 2019 |
| GRIN1 | 674 | Asn | Ile | 1.4 | 1.0 | M (1.5) | n.a. | M (1.7) | Increase | VCV001163324.5 | 29365063 | this study, Fry 2018 |
| GRIN1 | 688 | Ser | Tyr | M (0.48) | H (0.0016) | 0.73 | n.a. | H (0.13) | Decrease | VCV000981272.5 | 33122756 | this study, Skrenkova 2020 |
| GRIN1 | 732 | Asp | Glu | n.a. | n.a. | H (3.3) | 0.85 | n.a. | Increase | -- | 34884460 | Santos-Gómez 2021 |
| GRIN1 | 744 | Cys | Tyr | H (3.9) | M (1.8) | M (1.7) | H (4.8) | M (1.8) | Increase | VCV000539840.4 | 35393335 | Brock 2022 |
| GRIN1 | 794 | Arg | Gln | H (5.9) | M (1.6) | M (1.5) | H (3.6) | M (1.6) | Increase | VCV000435376.7 | 29365063 | this study, Fry 2018 |
| GRIN1 | 805 | Pro | Leu | H (6.1) | H (5.7) | 1.0 | H (3.4) | 1.3 | Increase | VCV000658023.10 | 34884460 | this study, Santos-Gómez 2021 |
| GRIN1 | 814 | Ala | Asp | H (0.34) | H (0.33) | 1.29 | H (0.43) | H (0.14) | Decrease | VCV000430264.2 | 34884460 | this study, Santos-Gómez 2021 |
| GRIN1 | 815 | Gly | Val | M (0.62) | 0.75 | H (3.3) | H (0.43) | H (0.052) | Decrease | -- | 30217972 | this study, Amin 2018 |
| GRIN1 | 815 | Gly | Arg | H (0.26) | M (0.5) | H (5.5) | H (0.48) | H (0.06) | Decrease | VCV001452432.3 | 27164704, 3021797[illegible] | this study, Lemke 2016; Amin 2018 |
| GRIN1 | 817 | Phe | Leu | M (0.49) | H (0.4) | 1.30 | M (0.52) | H (0.2) | Decrease | VCV000487504.3 | 30217972 | this study, Amin 2018 |
| GRIN1 | 818 | Met | Val | n.a. | n.a. | H (2.9) | 0.96 | n.a. | Increase | -- | 34884460 | Santos-Gómez 2021 |

| Gene | Residue Position | WT Amino Acid | Variant Amino Acid | Glutamate Potency | Glycine Potency | $Mg^{2+}$ IC50 | $\tau_{WEIGHT}$ | $P_{OPEN}$ | | ClinVar | PMID | Reference |
|---|---|---|---|---|---|---|---|---|---|---|---|---|
| GRIN2A | 184 | Ile | Ser | M (1.5) | 0.76 | 1.4 | H (4.8) | n.a. | Increase | VCV001325858.1 | 28611597 | this study, Sibarov 2017 |
| GRIN2A | 424 | Pro | His | 1.3 | 1.2 | H (2.8) | n.a. | n.a. | Increase | VCV000641065.27 | | this study |
| GRIN2A | 431 | Arg | Ser | 1.4 | M (1.9) | 1.3 | n.a. | n.a. | Increase | VCV001487019.6 | | this study |
| GRIN2A | 498 | Gly | Ser | M (0.51) | 0.72 | 1.3 | n.a. | n.a. | Decrease | VCV000930862.12 | | this study |
| GRIN2A | 614 | Asn | Ser | M (1.8) | M (1.6) | H (4.6) | n.a. | M (1.7) | Increase | VCV000224990.10 | 31429998 | Li 2019 |
| GRIN2A | 674 | His | Gln | 0.89 | 1.2 | H (3.5) | n.a. | n.a. | Increase | VCV001016692.7 | | this study |
| GRIN2A | 685 | Val | Gly | H (0.01) | 0.72 | n.a. | H (0.43) | n.a. | Decrease | VCV000205655.2 | 27839871 | this study, Swanger 2016 |
| GRIN2A | 733 | Ala | Thr | H (0.05) | 0.76 | 0.92 | n.a. | n.a. | Decrease | VCV000205658.13 | | this study |
| GRIN2A | 776 | Asp | Tyr | M (0.43) | 1.1 | 1.0 | n.a. | n.a. | Decrease | VCV000952584.9 | | this study |
| GRIN2A | 803 | Glu | Lys | H (0.24) | 0.72 | M (0.53) | n.a. | n.a. | Decrease | VCV001190362.2 | | this study |

| Gene | Residue Position | WT Amino Acid | Variant Amino Acid | Glutamate Potency | Glycine Potency | $Mg^{2+}$ IC50 | $\tau_{WEIGHT}$ | $P_{OPEN}$ | | ClinVar | PMID | Reference |
|---|---|---|---|---|---|---|---|---|---|---|---|---|
| GRIN2B | 499 | Gly | Val | H (0.14) | 0.75 | 1.2 | n.a. | 0.88 | **Decrease** | VCV001285574.1 | | this study |
| GRIN2B | 514 | Thr | Ala | H (0.011) | M (1.5) | 1.2 | n.a. | n.a. | **Decrease** | VCV000916599.1 | | this study |
| GRIN2B | 525 | Phe | Val | H (0.28) | 0.68 | 1.3 | n.a. | n.a. | **Decrease** | VCV000916600.1 | | this study |
| GRIN2B | 558 | Val | Ile | 1.2 | 0.9 | 0.9 | M (0.53) | H (0.06) | **Decrease** | VCV000374243.12 | 28377535, 2968179[illegible] | Platzer 2017; Vyklicky 2018 |
| GRIN2B | 687 | Pro | Leu | H (0.2) | 0.87 | 1.0 | n.a. | n.a. | **Decrease** | VCV000205711.2 | | this study |
| GRIN2B | 742 | Arg | Ile | H (5.9) | M (1.9) | 1.0 | n.a. | n.a. | **Increase** | VCV000523382.2 | | this study |
| GRIN2B | 751 | Ile | Leu | H (0.32) | H (0.3) | 1.3 | n.a. | n.a. | **Decrease** | VCV000245954.2 | | this study |
| GRIN2B | 754 | Gly | Arg | 1.3 | M (0.44) | 1.2 | n.a. | n.a. | **Decrease** | VCV000444285.23 | | this study |
| GRIN2B | 799 | Gly | Ser | H (17) | M (1.6) | 1.4 | n.a. | n.a. | **Increase** | VCV000373959.3 | | this study |
| GRIN2B | 819 | Ala | Thr | H (2.5) | H (6.7) | M (1.7) | n.a. | n.a. | **Increase** | VCV000916592.2 | 28377535 | Platzer 2017 |
| GRIN2B | 820 | Gly | Ala | 1.0 | 1.0 | 0.91 | H (0.31) | H (0.49) | **Decrease** | VCV000208643.20 | 30217972 | Amin 2018 |

Fold change was calcuated from ration of fitted parameters for mutant and WT controls recorded o the same day.
All calls were made by criteria of Myers et al., (2023). n.a.indicates data not available.
