## Supplemental Table S4 for "Distances from ligands as main predictive features for pathogenicity and functional effect of variants in NMDA receptors"

**Supplemental Table S4: Classification of enw variants reported here according to the criteria of Myers et al. (2023)**

| *GRIN* -variant | Glutamate Potency | Glycine Potency | $Mg^{2+}$ $IC_{50}$ | $P_{OPEN}$ | $\tau_{WEIGHT}$ | Surface Expression | Count (High, Mod) | Charge Transfer[a] | Classification |
|---|---|---|---|---|---|---|---|---|---|
| **1-K531R/2A** | H (0.46) | H (0.50) | 1.0 | H (0.38) | M (0.51) | 1.1 | 1, 3 | 0.27, 0.19 | Likely LoF |
| **1-Q556R/2A** | H (3.4) | H (3.1) | M (1.5) | H (2.3) | H (4.0) | 1.1 | 4, 1 | 7.1, 13 | Likely GoF |
| **1-P557R/2A** | H (10) | H (14) | 1.3 | H (0.12) | H (7.3) | M (0.38) | Conflict | 0.93, 2.8 | Possible GoF |
| **1-G638S/2A** | 1.3 | 1.3 | H (4.1) | 1.2 | 0.77 | M (0.60) | Conflict | 2.9, 5.3 | Possible GoF |
| **1-C744G/2A** | H (3.9) | 0.80 | 1.2 | M (1.6) | M (2.3) | 1.0 | 2, 1 | 5.3, 13 | Likely GoF |
| **1-A806E/2A** | H (10) | H (14) | 0.89 | 1.2 | H (8.7) | 1.2 | 3, 0 | 9.1, 14 | Likely GoF |
| **1-R844L/2A** | 1.3 | 1.4 | H (2.4) | 1.4 | 1.4 | 1.4 | subthreshold | 3.7, 3.6 | Possible GoF |
| **2A-R518C** | H (0.00053) | 0.90 | 0.81 | tstm | tstm | H (0.32) | tstm | tstm | Likely LoF* |
| **2A-T690M** | H (0.00050) | 0.86 | 1.2 | tstm | tstm | H (0.30) | tstm | tstm | Likely LoF* |
| **2A-I694T** | H (0.34) | 1.3 | 1.1 | 0.72 | 0.80 | 0.72 | 1, 0 | 0.51, 0.16 | Likely LoF |
| **2A-R695Q** | H (11) | 1.0 | 1.0 | 1.2 | H (3.6) | 0.77 | 2, 0 | 2.9, 14 | Likely GoF |
| **2A-A716D** | tstm | tstm | tstm | tstm | tstm | n.a. | tstm | tstm | Likely LoF* |
| **2A-K772E** | M (0.62) | 0.9 | M (0.63) | 0.78 | 0.84 | H (0.18) | 1, 2 | 0.083, 0.054 | Likely LoF |
| **2A-L812M** | H (8.7) | H (14) | M (2.0) | H (2.2) | H (9.4) | 0.85 | 4, 1 | 32, 49 | Likely GoF |
| **2A-M817V** | H (13) | H (6.7) | H (2.9) | H (5.81) | H (15) | 1.0 | 5, | 579, 860 | Likely GoF |
| **2A-A818V** | M (2.4) | M (2.4) | M (2.4) | H (4.1) | M (1.7) | M (1.6) | 1, 5 | 24, 43 | Likely GoF |
| **2B-G543R** | H (5.4) | H (3.4) | 1.2 | M (0.65) | H (3.3) | 1.1 | Conflict | 1.8, 3.7 | Possible GoF |
| **2B-V558I** | 1.2 | 0.92 | 0.86 | 0.06 | 0.53 | 1.0 | 1, 1 | 0.03, 0.07 | Likely LoF |
| **2B-W559C** | tstm | tstm | tstm | tstm | tstm | 0.74 | tstm | tstm | Likely LoF* |
| **2B-W559R** | tstm | tstm | tstm | tstm | tstm | 0.81 | tstm | tstm | Likely LoF* |
| **2B-N616I** | H (5.1) | H (5.8) | H (1641) | 0.75 | H (3.2) | 0.82 | 4, 0 | 95, 191 | Likely GoF |
| **2B-N616S** | 1.3 | 1.3 | H (1778) | H (0.28) | M (1.6) | 0.83 | Conflict | 11, 9.3 | Possible GoF |
| **2B-M654I** | H (20) | H (9.9) | M (1.8) | 0.86 | H (9.8) | 0.67 | 3, 1 | 6.1, 11 | Likely GoF |
| **2B-M654V** | H (21) | H (3.9) | M (1.8) | H (3.8) | H (2.1) | H (0.25) | Conflict | 2.1, 18 | Possible GoF |
| **2B-T691A** | H (0.00073) | 0.94 | 1.1 | tstm | tstm | 0.96 | tstm | tstm | Likely LoF* |
| **2B-T691I** | H (0.0011) | 0.81 | 1.1 | tstm | tstm | 1.2 | tstm | tstm | Likely LoF* |
| **2B-R696C** | H (4.6) | 0.96 | 0.99 | 0.82 | H (2.7) | 0.96 | 2, 0 | 2.8, 5.9 | Likely GoF |
| **2B-A717E** | tstm | tstm | tstm | tstm | tstm | M (0.41) | tstm | tstm | Likely LoF* |
| **2B-C746W** | 1.0 | H (0.12) | 0.71 | 0.68 | 0.86 | M (0.47) | 2, 0 | 0.078, 0.095 | Likely LoF |
| **2B-L797F** | H (0.041) | M (0.65) | 0.93 | 1.4 | H (0.054) | 0.89 | 2, 1 | 0.062, 0.019 | Likely LoF |
| **2B-L813V** | H (5.4) | H (13) | 0.89 | 1.1 | M (1.9) | 0.83 | 2, 1 | 1,2, 4.6 | Likely GoF |
| **2B-M818R** | tstm | tstm | tstm | tstm | tstm | M (0.39) | tstm | tstm | Likely LoF* |
| **2B-M818T** | M (2.5) | M (2.2) | 0.68 | H (3.4) | H (2.4) | M (0.63) | Conflict | 3.5, 4.3 | Possible GoF |
| **2B-G820A** | 1.0 | 1.0 | 0.91 | H (0.49) | H (0.31) | 0.84 | 2,0 | 0.16, 0.53 | Likely LoF |

Fold change was calcuated from ration of fitted parameters for mutant and WT controls recorded o the same day.

All calls were made by criteria of Myers et al., (2023). N.a.a indicates data not available.

tstm indicates that the current response was too small to measure despite measureable surface expression
